# Physical Fitness and Body Composition in Transgender vs. Cisgender Individuals: A Systematic Review and Meta-Analysis

**DOI:** 10.1101/2025.05.05.25326994

**Authors:** Sofia Mendes Sieczkowska, Bruna Caruso Mazzolani, Danilo Reis Coimbra, Igor Longobardi, Andresa Rossilho Casale, José Davi F.V.M.P. da Hora, Hamilton Roschel, Bruno Gualano

## Abstract

**Objective:** To compare fitness and body composition between transgender and cisgender individuals.

**Design:** Systematic review and meta-analysis.

**Data sources:** PubMed, Web of Science, Embase, and SportDiscus databases were searched in June 2024, supplemented by manual citation reviews.

**Eligibility criteria:** Inclusion criteria comprised studies of transgender individuals comparing physical fitness/body composition pre-to-post gender-affirming hormone therapy (GAHT) or vs. cisgender controls, with quantitative outcomes reported.

**Results:** Fifty-one studies (6,434 participants) were analyzed. Transgender women (TW) exhibited comparable fat mass to cisgender women (CW), but higher than cisgender men (CM). TW showed greater lean mass than CW, but lower than CM. Upper- and lower-body strength were similar between TW and CW, but lower than CM. TW and CW had similar VO_2_ peak, but TW exhibited lower values than CM. Transgender men (TM) exhibited similar fat mass to CW, but higher than CM. TM showed higher lean mass than CW, but lower than CM. Upper-body strength was higher in TM than CW, but lower than CM. GAHT in TW increased fat mass, reduced lean mass and upper-body strength, with no differences in lower-body strength over 1–3 years. TM demonstrated reduced fat mass and increased lean mass, upper- and lower-body strength post-GAHT. Risk of bias was moderate for most studies, with limited observations for specific outcomes (e.g., VO_2_ peak).

**Conclusion:** While TW exhibited higher lean mass than CW, their physical fitness was comparable. Current evidence is limited but does not justify blanket bans based on assumptions of inherent athletic advantages for TW over CW.

**WHAT IS ALREADY KNOWN ON THIS TOPIC:**

- The inclusion of transgender women in female sports categories remains a highly contentious subject.
- Existing studies suggest that gender-affirming hormone therapy (GAHT) alters body composition in transgender individuals, but evidence on functional performance outcomes (e.g., strength, endurance) remains inconsistent.
- Policies advocating blanket bans on transgender women (TW) in female sports often cite residual advantages from prior testosterone exposure, despite limited empirical support for sustained performance disparities post-GAHT.

**WHAT THIS STUDY ADDS?:**

- This systematic review and meta-analysis synthesizes data from 51 studies (6,434 participants), demonstrating that, while TW showed higher absolute lean mass than cisgender women (CW), there are no significant differences in upper/lower-body strength or VO□peak after 1–3 years of GAHT.
- These findings challenge the validity of blanket bans predicated on assumptions of inherent athletic superiority of TW over CW.
- Transgender men (TM) exhibit body composition and strength metrics intermediate between CW and cisgender men (CM) post-GAHT.
- Critical research gaps are identified, including a lack of long-term GAHT data, underrepresentation of transgender athletes, and inconsistent controls for confounders (e.g., training history, puberty blocking). Future studies must prioritize sport-specific performance metrics and longitudinal designs to inform equitable policies.

## Introduction

The question of whether transgender women (TW) should be permitted to compete in female sports, and under what conditions, remains a subject of intense debate. The rationale for sex-segregated competition is rooted in ensuring equitable opportunities for cisgender women (CW), prompting proposals that transgender (or intersex) athletes should be included only if their participation does not disproportionately disrupt competitive fairness [1, 2, 3].

However, empirical evidence challenges initial concerns that TW would dominate women’s sports, largely due to the physiological effects of testosterone suppression therapy [1]. In fact, TW remain underrepresented in elite athletics. For example, Laurel Hubbard, the first openly transgender woman to compete in the Tokio Olympic Games (2021), participated in weightlifting but did not advance to medal contention, underlining the lack of dominance by transgender athletes in practice.

The International Olympic Committee (IOC) recently established a framework prioritizing fairness, inclusion, and nondiscrimination for athletes with diverse gender identities and sex variations [2]. This approach rejects blanket bans based on gender identity, advocating instead for sport-specific eligibility criteria informed by evidence. Critics, however, argue that the framework relies on insufficiently developed research and impractical case-by-case assessments, potentially compromising protections for cisgender female athletes. Lundberg et al. (2024) [3] contend that the IOC’s “no presumption of advantage” principle overlooks studies suggesting that transgender women retain muscle mass, strength, and other physical advantages over cisgender women even after testosterone suppression. Their argument hinges on the well-established physiological disparities between cisgender males and females, which confer inherent athletic advantages to males. Nonetheless, systematic reviews comparing transgender women (post-hormone therapy) and cisgender women report inconsistent findings regarding performance and physical differences, highlighting the need for further research [1, 4].

To shed light on this topic, this systematic review with meta-analysis evaluates the current literature on the physical fitness and body composition of transgender individuals (i.e., TW and transgender men [TM]) relative to cisgender ones (i.e., CW and cisgender men [CM]). By synthesizing available data, we aim to guide decision-makers and stakeholders in developing equitable, scientifically grounded policies for transgender participation in sports.

## Methods

This systematic review was registered in PROSPERO (CRD42024562210) and follow the guidelines of the PRISMA 2020 statement [5].

### Search Strategy

For comprehensive coverage of worldwide scientific production, the search was conducted in the following electronic databases: PubMed, Web of Science, Embase, and SportDiscus during the first week of June 2024. The search strategy used descriptors related to the population and outcomes as follows: Population (Transgender OR Transexual OR Transgender Person OR Transsexualism OR Transgenderism OR “Transgender people” OR “gender reassignment procedure” OR “gender reassignment surgery” OR “gender change procedure” OR “Gender-affirming Treatment” OR transwoman OR transmen OR “gender affirming hormone therapy (GAHT)” OR transsexual OR “cross-sex hormone therapy” OR “trans people”) and Outcomes (physical fitness and body composition terms, see table S1 supplementary material). To select search descriptors, MeSH terms (Medical Subject Headings) were used. Additionally, a manual search of the references in selected studies were conducted to identify studies for inclusion. The search process was carried out independently by two researchers. In case of disagreement, a third reviewer was consulted.

### Eligibility for Study Selection

All articles identified in the search were screened by two independent members of the research team using a 3-stage strategy: 1) Title and abstract screening, and 2) Full text review and 3) Conflict resolution / consensus phase. Any discrepancies were resolved through discussion, or third-party mediation, if required. The selection was made using the software Rayyan QCRI [6].

Study eligibility was based on the PECO criteria, described below:

Population: Individuals identifying as transgender.

Exposure: Gender-affirming hormone therapy.

Comparison: Before and after gender-affirming hormone therapy, and with cisgender individuals.

Outcomes: Measures of physical fitness and body composition.

No language restrictions were applied, and there were no restrictions on publication dates.

### Data Extraction Process

A data extraction table was created and completed to gather key information from the selected studies, including author, study population, sample size, type of hormonal therapy, outcomes, and main findings.

### Risk of bias

The risk of bias analysis was conducted using the 20-item AXIS (Appraisal tool for cross-sectional studies) [7], ROBINS-I (Risk Of Bias In Non-randomized Studies - of Interventions) [8] or ROB2 (Risk Of Bias 2) [9], depending on study design.

AXIS was used to assess the risk of bias in cross-sectional studies. It consists of 20 items, each rated as “Yes,” “No,” or “Don’t Know.” Items 7 (“Were measures undertaken to address and categorize non-responders?”) and 14 (“If appropriate, was information about non-responders described?”) were marked as “Inapplicable” when the response rate was reported as 100%. As AXIS lacks a standardized scoring system, we adopted a method used in previous studies [10–12], assigning scores of 0 or 1 to each item to calculate an overall quality score. Specifically, for items 13 (“Does the response rate raise concerns about non-response bias?”) and 19 (“Were there any funding sources or conflicts of interest that may affect the authors’ interpretation of the results?”), a response of “No” was scored as 1, and “Yes” or “Don’t Know” as 0. For items 7 and 14, “Yes” or “Inapplicable” were scored as 1, and “No” or “Don’t Know” as 0. For all other items, “Yes” was scored as 1, and “No” or “Don’t Know” as 0. Total scores were calculated by summing the individual item scores. Following recommendations [13], studies were classified as high quality (scores of 14–20; 70–100%), fair quality (scores of 12–13; 60–69.9%), or low quality (scores of 0–11; 0–59.9%).

ROBINS-I was used to assess the risk of bias in cohorts and quasi-experimental studies, and includes seven domains of bias: bias due to confounding, bias in participants’ selection, bias in interventions classification, bias due to deviations from intended interventions, bias due to missing data, bias in outcomes measurement, and bias in reported result selection. Each domain is assessed through signaling questions to provide a comprehensive analysis of potential biases within the study. Each domain is assessed through signaling questions to provide a comprehensive analysis of potential biases within the study.

ROB2 was used to assess the risk of bias in randomized controlled trials (RCTs) and consists of five domains: bias arising from the randomization process, bias due to deviations from intended interventions, bias due to missing outcome data, bias outcome measurement, and bias in reported result selection. Similar to ROBINS-I, it also incorporates a set of signaling questions to determine whether each domain poses a high, low, or unclear risk of bias.

Two independent members conducted this analysis. Any discrepancies were resolved through discussion or third-party mediation, if required.

### Statistical analysis

The metacont function in RStudio was used to perform a comprehensive statistical analysis of the effects of hormone therapy on transgender individuals. This analysis involved aggregating data from studies comparing baseline and post-hormone therapy measures, focusing on the impact of hormone therapy on key physical fitness and body composition parameters. A random-effects model was employed to account for variability both within and between studies, enabling a robust estimation of the overall effect of hormone therapy on physical fitness outcomes in transgender individuals. The analyses were conducted only when there were at least three studies for each subgroup.

In addition, the metacont function was also used to compare the effects of gender-affirming hormone therapy between transgender and cisgender individuals. This analysis synthesized data from multiple studies examining the same physical fitness and body composition variables, aiming to quantitatively assess the magnitude and direction of treatment effects in transgender in comparison to cisgender individuals. This approach provided valuable insights into the impact of gender-affirming hormone therapy on physical fitness and body composition parameters. The analyses were conducted only when there were at least three studies for each subgroup. The entire analysis was conducted using RStudio (version 4.4.0) with the meta statistical package [14].

## Results

The database search identified 1,705 studies. Two studies published later were included after checking the citations of the included articles and other sources (Google Scholar, social media). After removing duplicates (n = 638), 1,067 publications were screened for inclusion. Of these, 595 were excluded based on title review and 284 after abstract review. The remaining 188 papers were selected for full-text reading, from which three were excluded due to the population, 48 for outcome, and 86 for “wrong publication type”. Therefore, 51 studies were included in the review [15–65] and 42 in the meta-analysis (Figure S1).

### Methodological Characteristics of the Studies

The analysis included a total of 6,434 individuals: 2,926 TW, 2,309 TM, 551 CW, and 648 CM. Participants’ mean age ranged from 14 to 41 years. Of the 51 studies reviewed, 44 focused on adults, while seven involved adolescents. Regarding study design, 22 were prospective cohorts, nine retrospective cohorts, 16 cross-sectional studies, three randomized controlled trials, and one quasi-experimental studies. Only 15 studies incorporated any form of physical activity (PA) assessment. Among these, two exclusively recruited amateur athletes, and another reported including only sedentary individuals.

**Table 1.**
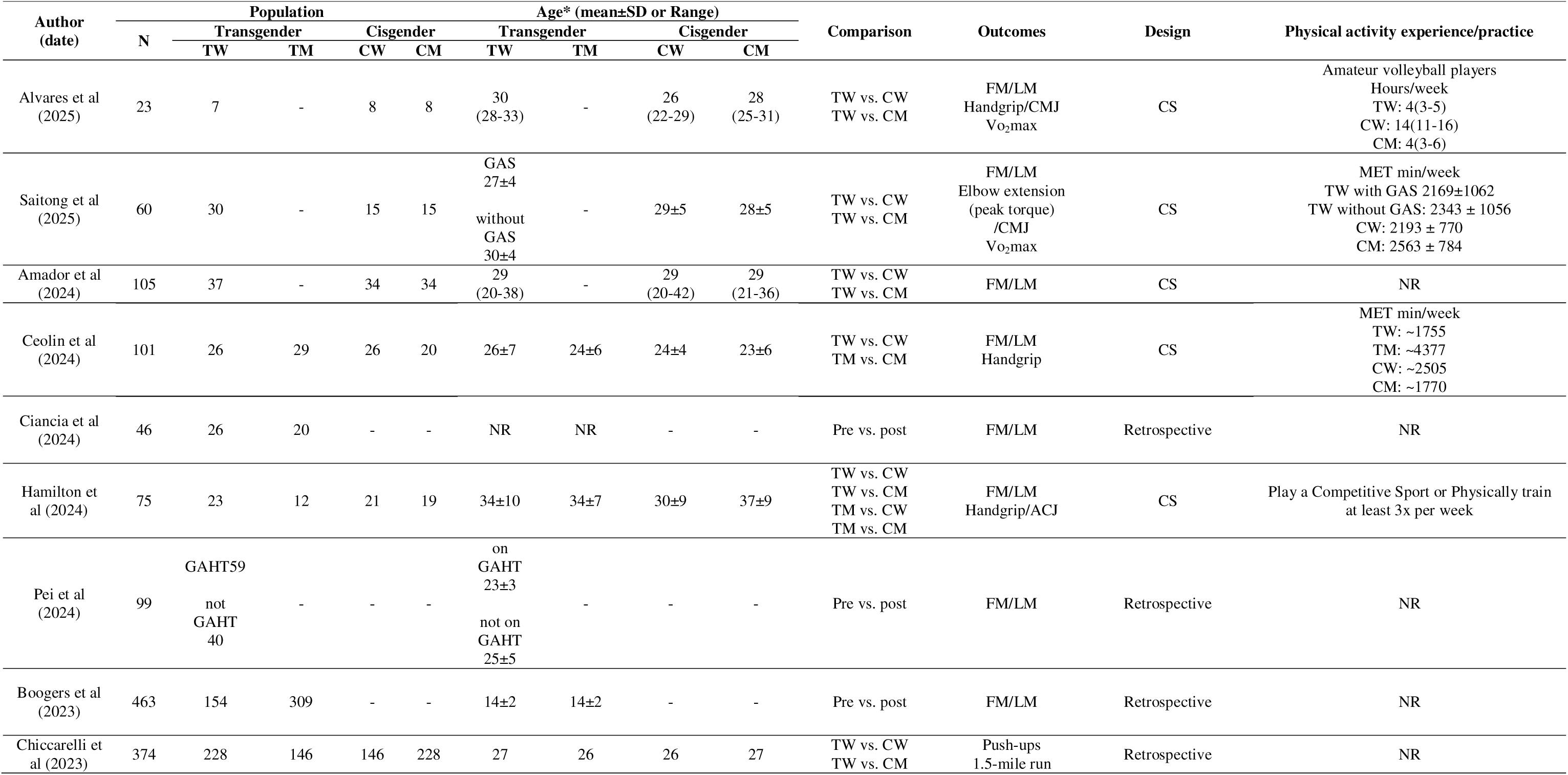
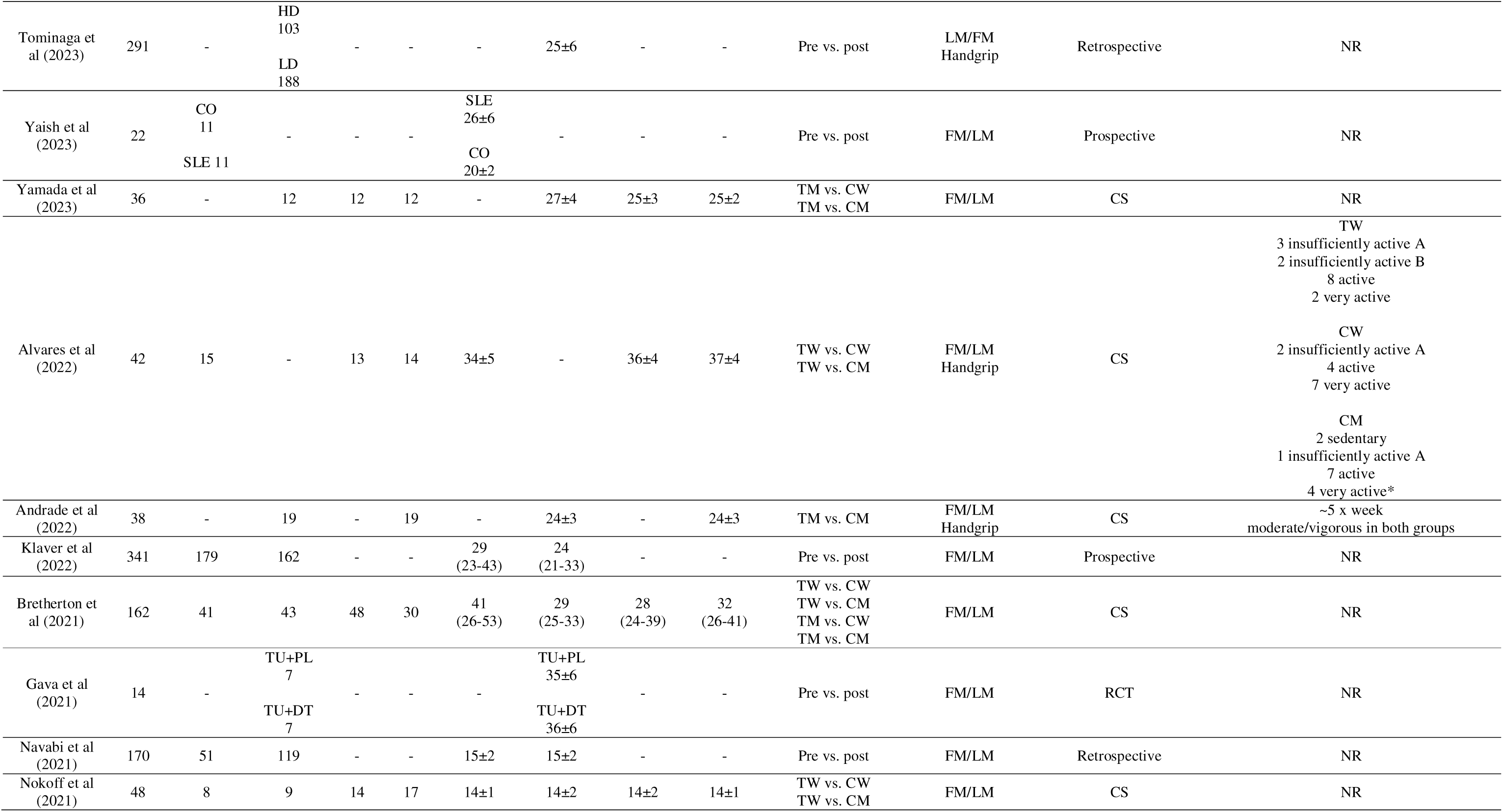
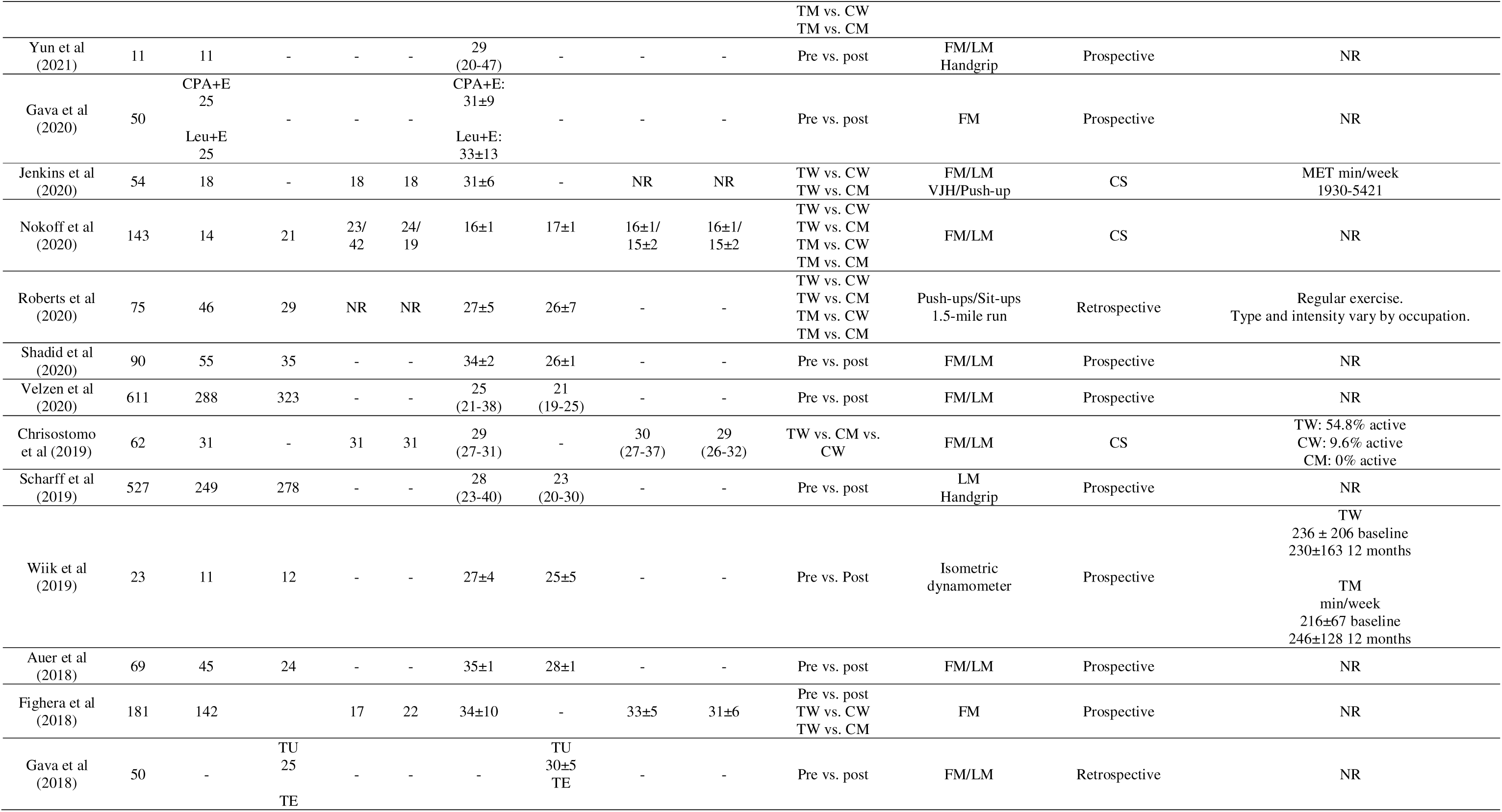
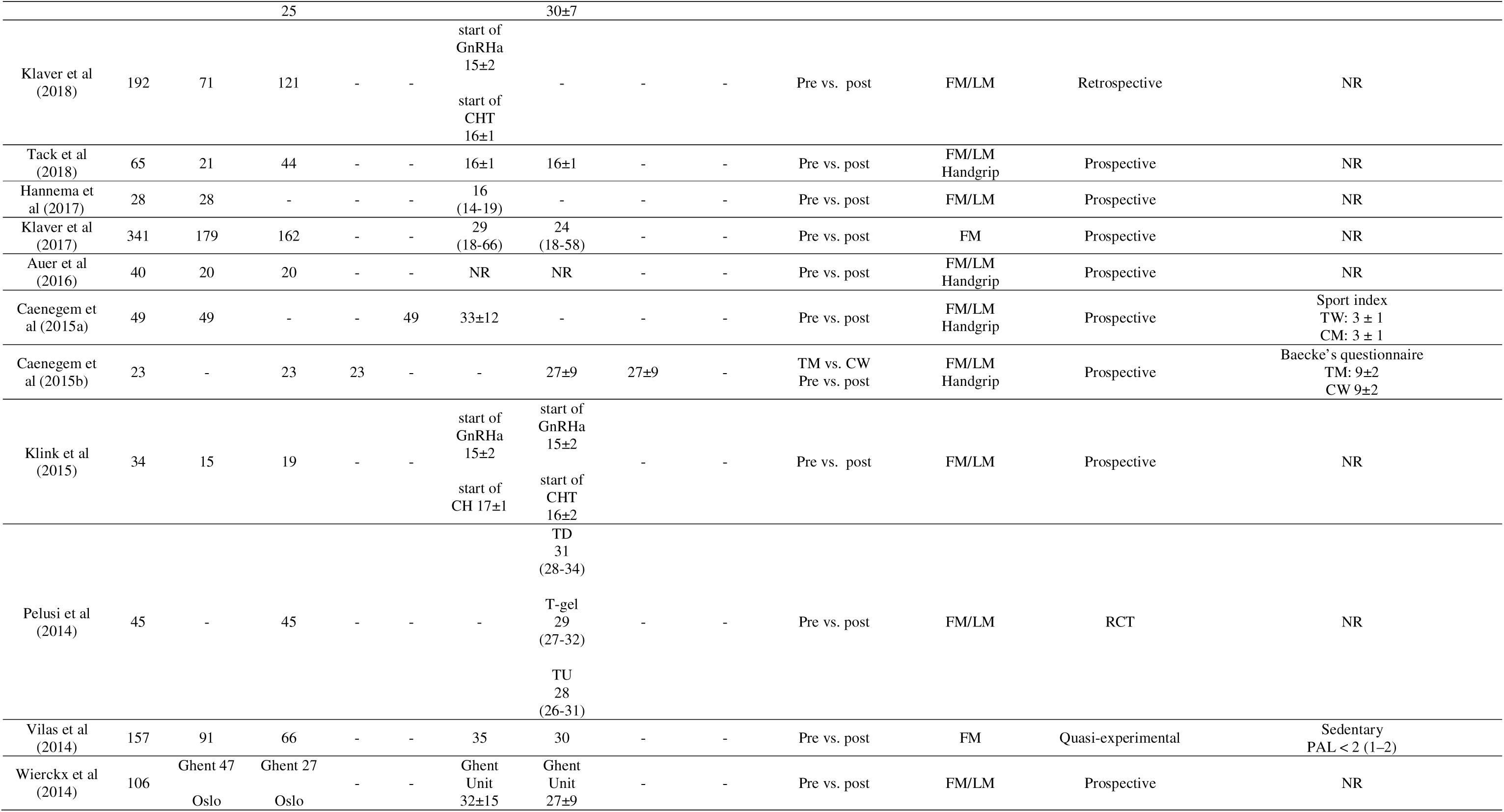
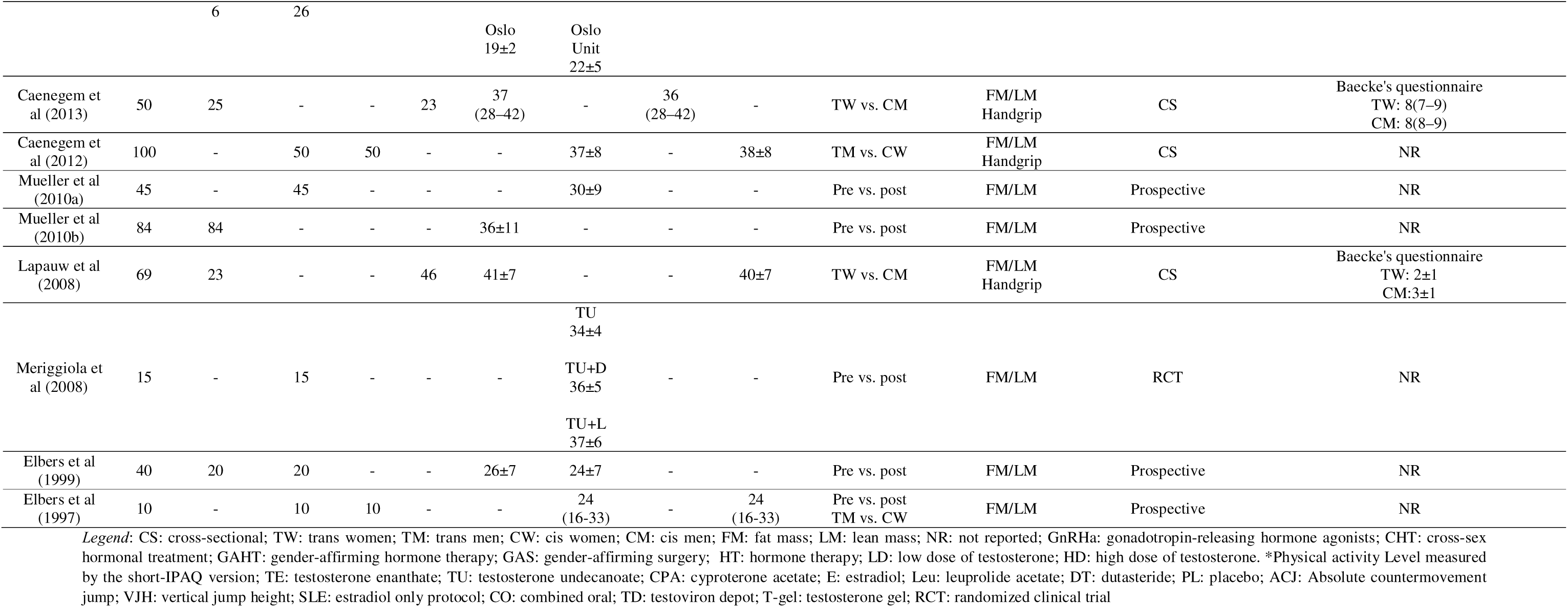
Methodological characteristics of the studies included.

Methods for assessing PA levels varied significantly: three studies used weekly METs (metabolic equivalents); three applied the Baecke questionnaire (mean scores); two reported the percentage or number of “active” participants; two documented weekly exercise frequency; two measured weekly exercise duration; one used the Sport Index (mean scores); one simply noted “regular physical activity” without further detail, and one reported including only sedentary individuals (PAL < 2.0).

Reviewed studies provided details on various hormonal therapies and related findings (Table 2). Among reported treatments, GAHT was used in 49 studies, either alone or in combination with: Antiandrogens (18 studies, exclusively for TW), GnRH agonists (GnRHa) (six studies). GnRHa alone was used in two studies.

The cross-sex hormone therapies included estrogen formulations (estradiol valerate, 17β-estradiol, estradiol gel, and transdermal estradiol); testosterone formulations (testosterone enanthate, testosterone undecanoate [administered intramuscular, at low or high dose]); antiandrogens (cyproterone acetate, spironolactone, and finasteride); GnRH agonists (triptorelin, leuprolide acetate or goserelin acetate).

Therapy duration varied widely, ranging from 3 months to 14 years, with most studies reporting following participants for 1 to 3 years of therapy. Also, of the 51 studies included, 11 included participants who had undergone gender-affirming surgery and only six reported the use of puberty suppression.

In 15 cross-sectional studies, information regarding naïve status to hormone therapy, adverse effects, and dropouts were deemed less relevant due to the study design. Therefore, of the remaining 36 studies included, 23 exclusively enrolled hormone-naïve individuals (one focused solely on naïve TW), five included non-naïve participants and eight did not report this data. Regarding adverse effects, 27 studies did not report, eight observed none and one study reported several adverse events in both TM (muscle/joint pain, mild hypertension, reduced fasting insulin, androgenic alopecia) and TW (depression, elevated prolactin, galactorrhea, transient liver enzyme elevations, hypertension, increased fasting insulin, and skin irritation). Moreover, 20 studies lacked dropout data, 11 reported none and four had significant attrition (≥20 participants) and one had minimal dropouts (n=2).

### Transgender compared with cisgender individuals

#### Fat Mass

Of the 22 studies reporting this comparison, 20 were included in the meta-analysis. Two studies were excluded because they either lacked extractable data (due to graphical format) or did not report mean/median values (or other measures of central tendency) for the outcome of interest.

In the comparison between TW and CW, the average duration of cross-sex hormone therapy for TW was 2.82 ± 2.09 (range: 1-7) years. No significant differences were observed between groups [SMD: -0.22, 95% CI (-0.62; 0.19)] (Figure 1).

**Figure 1.**
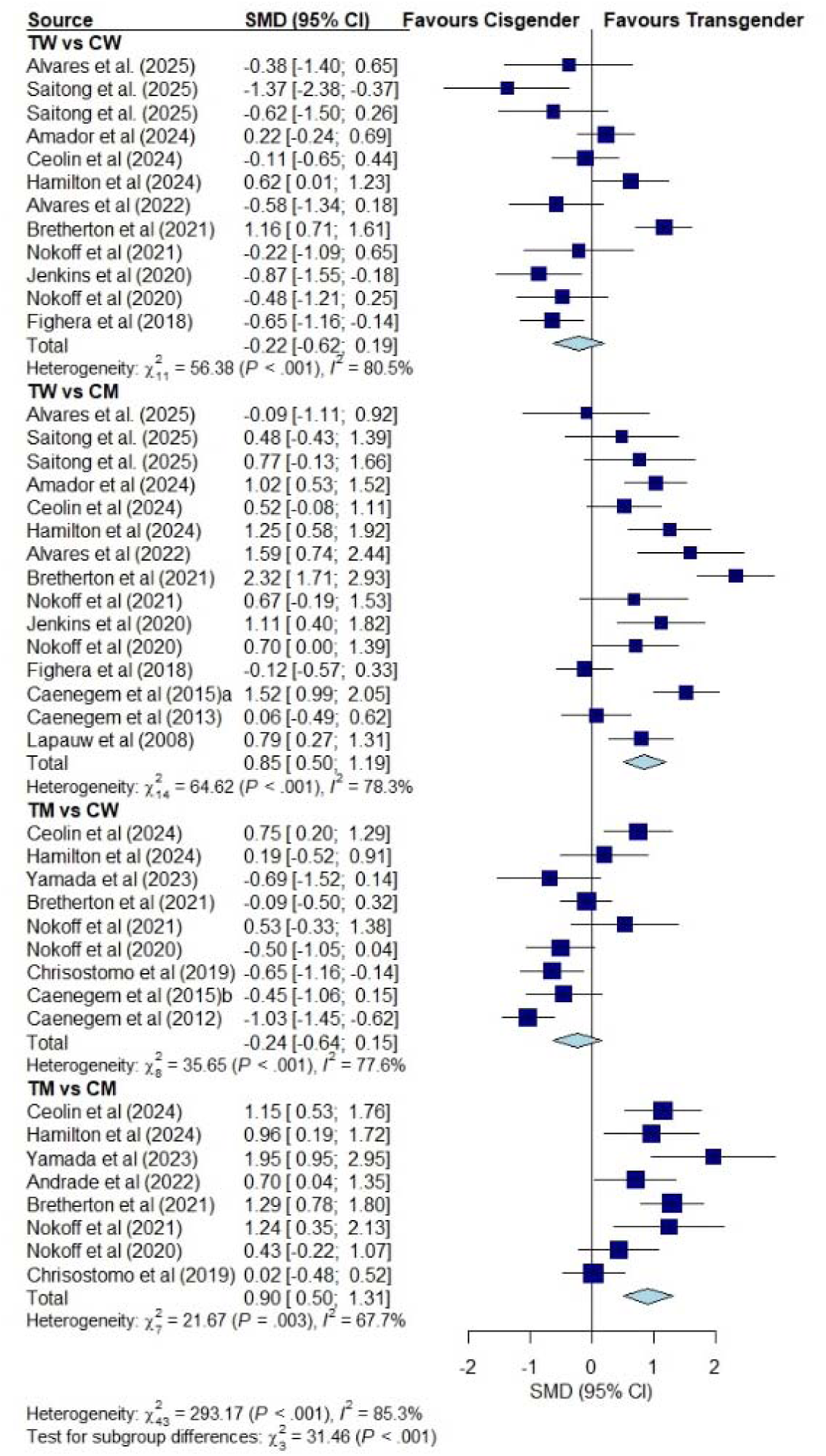
Forest plot: Fat mass in transgender women vs. cisgender women, transgender women vs. cisgender men, transgender men vs. cisgender women, and transgender men vs. cisgender men. Legend: TW: transgender women; TM: transgender men; CW: cisgender women; CM: cisgender men; SMD: standard mean difference; CI: confidence interval.

Average duration of cross-sex hormone therapy for TW was 3.25 ± 2.49 (range: 1-8) years when comparing with CM, with TW presenting higher fat mass [SMD: 0.85, 95% CI (0.50; 1.19)] (Figure 1).

Average duration of cross-sex hormone therapy for TM was 3.13 ± 3.09 (range: 1-10) years in the comparison with CW. No significant differences were observed between groups [SMD: -0.24, 95% CI (-0.64; 0.15)] (Figure 1).

Dataset used to compare TM and CM showed an average duration of cross-sex hormone therapy of 4.0 ± 4.62 (range: 1-14) years for TM. TM had a higher fat mass than CM [SMD: 0.90, 95% CI (0.50; 1.31)] (Figure 1).

#### Lean Mass

Of the 21 studies reporting this comparison, 18 were included in the meta-analysis. Three studies were excluded due to lack of extractable data (due to graphical format) or not reporting mean/median values (or other measures of central tendency) for the outcome of interest.

Cross-sex hormone therapy for TW was 2.80 ± 2.20 (range: 1-7) years on average when comparing with CW, and TW had a higher lean mass [SMD: 0.98, 95% CI (0.03; 1.93)] (Figure 2).

**Figure 2.**
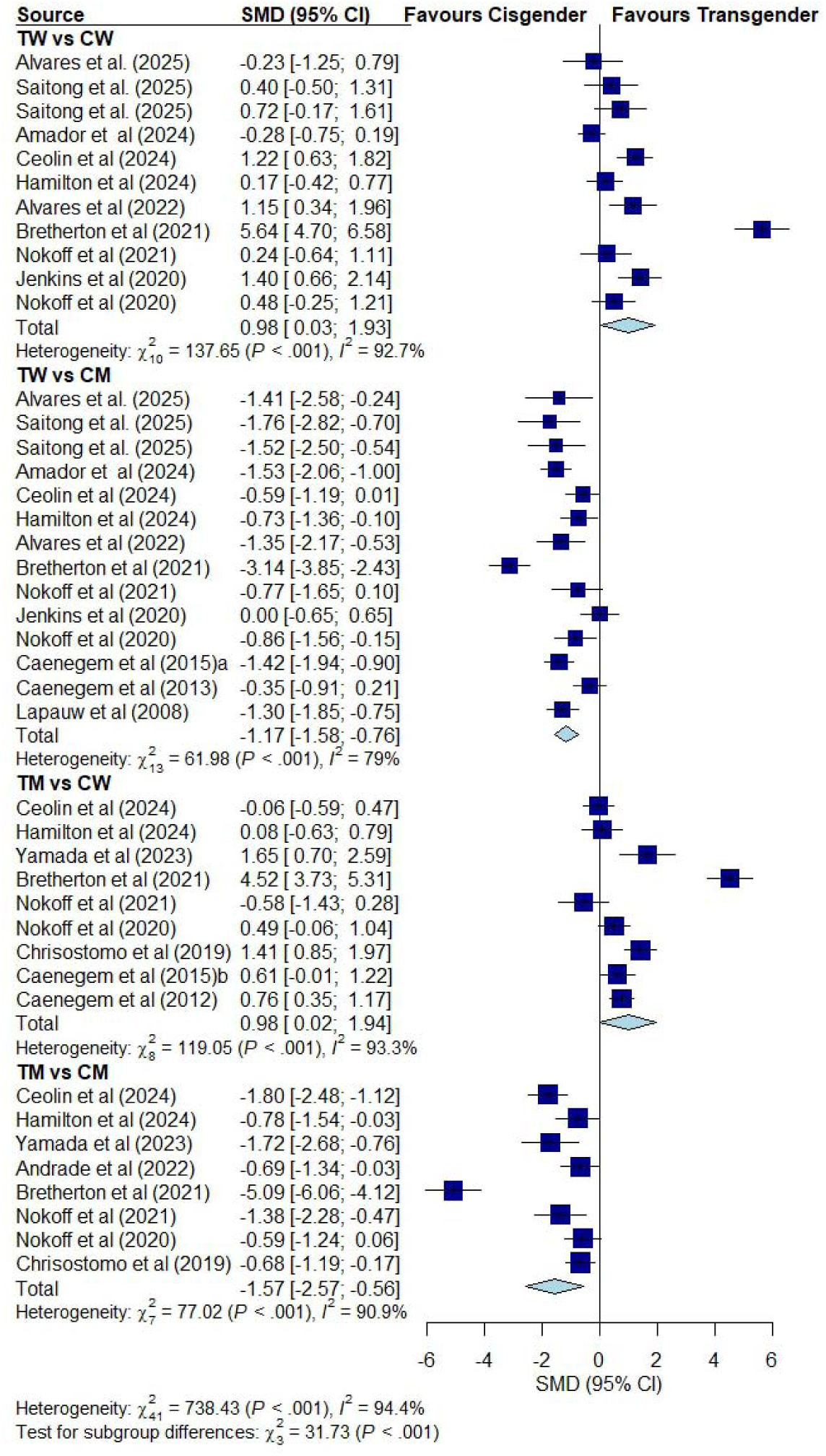
Forest plot: Lean mass in transgender women vs. cisgender women, transgender women vs. cisgender men, transgender men vs. cisgender women, and transgender men vs. cisgender men. Legend: TW: transgender women; TM: transgender men; CW: cisgender women; CM: cisgender men; SMD: standard mean difference; CI: confidence interval.

In the comparison between TW and CM, the average duration of cross-sex hormone therapy was 3.67 ± 2.84 (range: 1-8) years and TW exhibited a lower lean mass [SMD: -1.17, 95% CI (-1.58; -0.76)] (Figure 2).

Average duration of cross-sex hormone therapy for TM was 3.89 ± 3.69 (range: 1-10) years in the comparison with CW and TM exhibited a higher lean mass [SMD: 0.98, 95% CI (0.02; 1.94)] (Figure 2).

TM cross-sex hormone therapy duration average was 4.0 ± 4.62 (range: 1-14) years when comparing with CM, with TM exhibiting lower lean mass [SMD: -1.57, 95% CI (-2.57; -0.56)] (Figure 2).

#### Upper-body strength

All 12 studies reporting this outcome were included in the meta-analysis.

TW average duration of cross-sex hormone therapy was 3.17 ± 2.64 (range: 1-7) years. No significant differences were observed between groups when compared with CW [SMD: 0.41, 95% CI (-0.10; 0.92)] (Figure 3).

**Figure 3.**
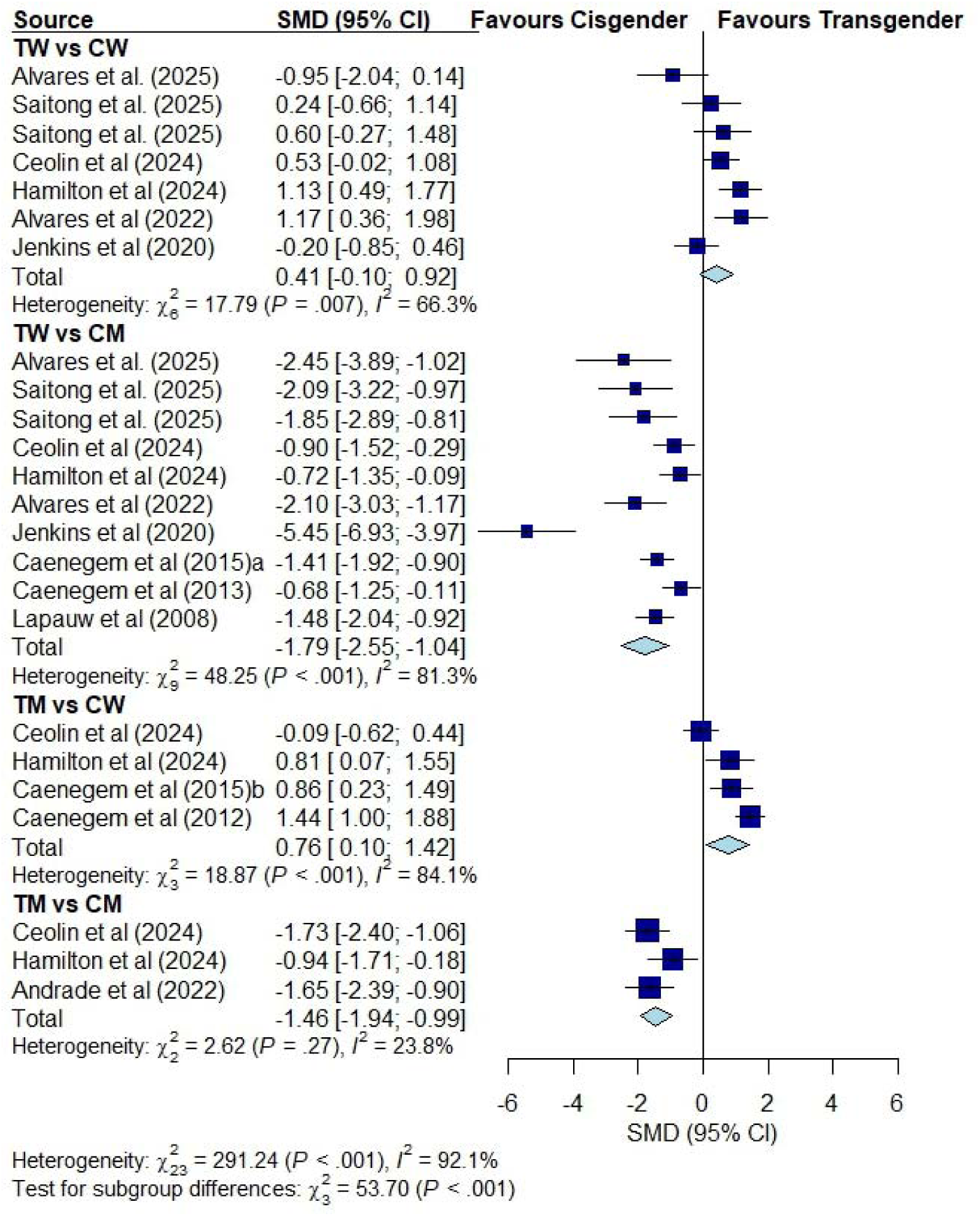
Forest plot: Upper-body strength in transgender women vs. cisgender women, transgender women vs. cisgender men, transgender men vs. cisgender women, and transgender men vs. cisgender men. Legend: TW: transgender women; TM: transgender men; CW: cisgender women; CM: cisgender men; SMD: standard mean difference; CI: confidence interval.

When comparing TW with CM, average duration of cross-sex hormone therapy was 3.86 ± 3.02 (range: 1-8) years. TW had a lower upper-body strength [SMD: -1.79, 95% CI (-2.55; -1.04)] (Figure 3).

In the comparison between TM and CW, the average duration of cross-sex hormone therapy for TM was 5.0 ± 4.58 (range: 1-10) years. TM had a higher upper-body strength than CW [SMD: 0.76, 95% CI (0.10; 1.42)] (Figure 3).

Average duration of cross-sex hormone therapy for TM was 9.0 ± 7.07 (range: 1-14) years when comparing with CM, and TM had a lower upper-body strength [SMD: -1.46, 95% CI (-1.94; -0.99)] (Figure 3).

#### Lower-body strength

Four studies reported this outcome and were included in the meta-analysis.

In the comparison between TW and CW, the average duration of cross-sex hormone therapy was 2.75 ± 2.87 (range: 1-7) years. No significant differences were observed between groups [SMD: 0.05, 95% CI (-0.74; 0.83)] (Figure 4).

**Figure 4.**
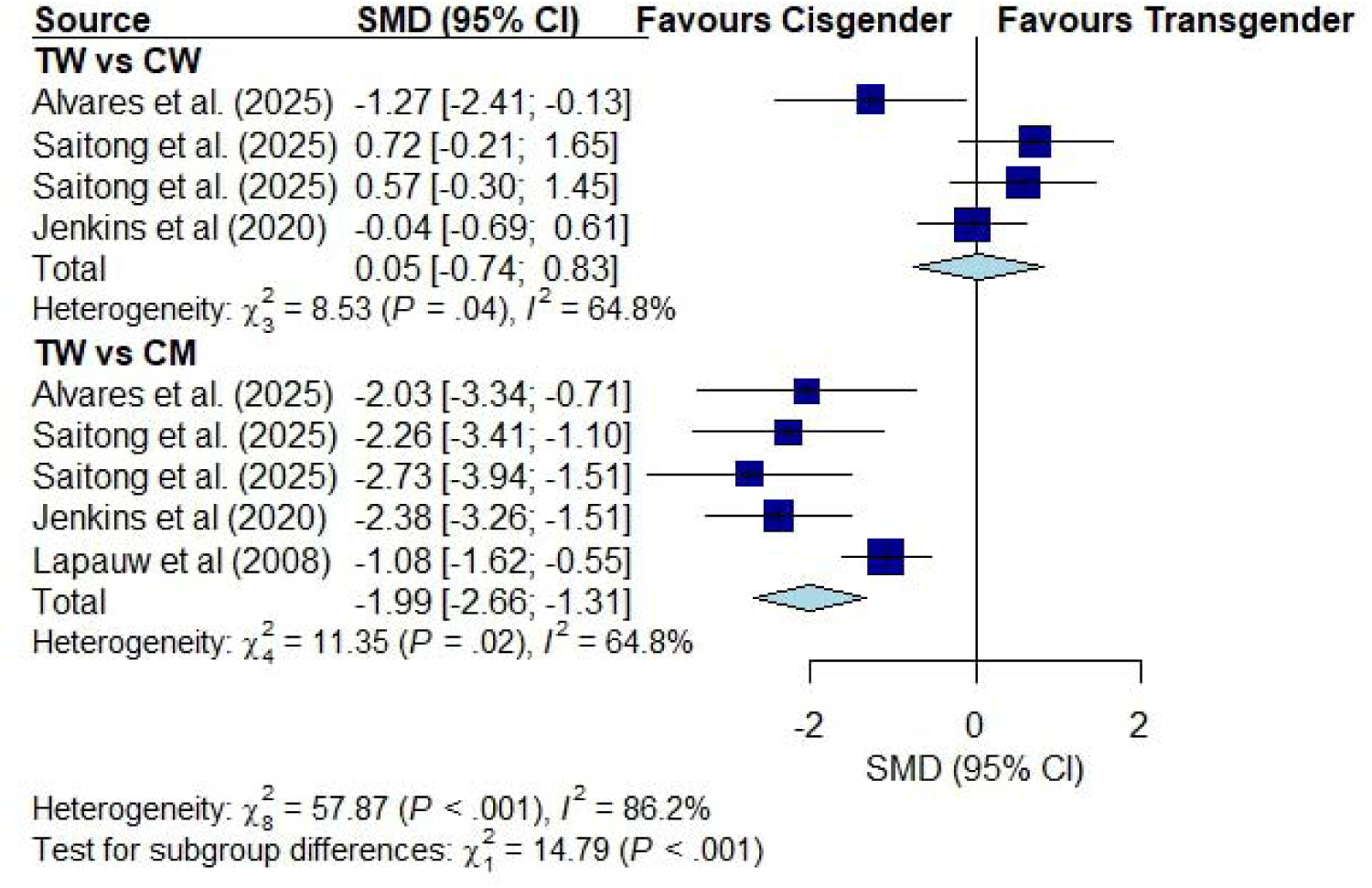
Forest plot: Lower-body strength in transgender women vs. cisgender women and transgender women vs. cisgender men. Legend: TW: transgender women; TM: transgender men; CW: cisgender women; CM: cisgender men; SMD: standard mean difference; CI: confidence interval.

Average duration of cross-sex hormone therapy for TW was 3.80 ± 3.42 (range: 1-8) years when comparing with CM, with TW exhibiting lower lower-body strength [SMD: -1.99, 95% CI (-2.66; -1.31)] (Figure 4).

#### VO_2_ peak

All three studies reporting this outcome were included in the meta-analysis. For VO_2_ peak, comparisons were only possible with TW, and average duration of cross-sex hormone therapy was 2.75 ± 2.87 (range: 1-7) years. No significant differences were observed between TW and CW [SMD: 0.07, 95% CI (-0.67; 0.82)] (Figure 5). TW exhibited lower VO_2_ peak when compared to CM [SMD: -1.61, 95% CI (-2.69; -0.54)] (Figure 5).

**Figure 5.**
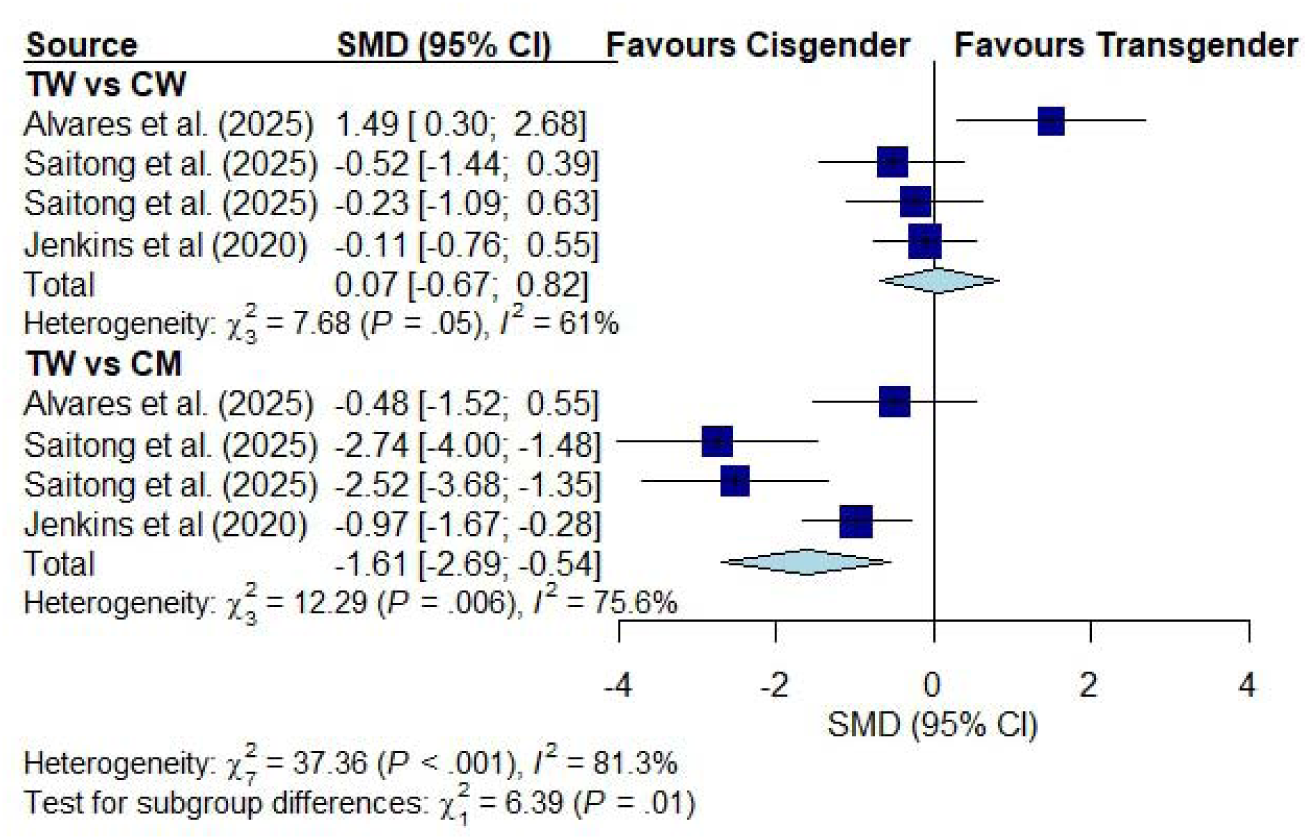
Forest plot: VO□peak in transgender women vs. cisgender women and transgender women vs. cisgender men. Legend: TW: transgender women; TM: transgender men; CW: cisgender women; CM: cisgender men; SMD: standard mean difference; CI: confidence interval.

### Effects of cross-sex hormone therapy in transgender individuals

#### Fat mass

Twenty-three out of the 28 studies reporting this outcome were included in the meta-analysis. Five studies were excluded due to lack of extractable data (due to a graphical format) or lack of report on the duration of cross-sex hormone therapy.

First, we conducted the analysis regardless of whether the individuals were naïve or had undergone puberty suppression.

In TW, an increase in body fat mass was observed after one year of cross-sex hormone therapy compared to baseline [SMD: 0.44, 95% CI (0.27; 0.61)] (Figure S2). A similar effect was observed in studies with follow-up between one and two years [SMD: 0.59, 95% CI (0.22; 0.96)] and in those with a three-year follow-up [SMD: 0.70, 95% CI (0.19; 1.21)] (Figure S2).

When considering studies with hormone-naïve TW, the increase in body fat mass persisted up to one year of therapy compared to baseline [SMD: 0.50, 95% CI (0.36; 0.65)], between one and two years [SMD: 0.89, 95% CI (0.59; 1.18)], and after three years [SMD: 1.29, 95% CI (0.86; 1.73)] (Figure S3).

In TM, no increase in body fat mass was observed after one year of cross-sex hormone therapy compared to baseline [SMD: -0.15, 95% CI (-0.36; 0.06)] (Figure S4). The same result was observed between one and two years [SMD: -0.13, 95% CI (-0.48; 0.23)] and after three years [SMD: -0.24, 95% CI (-0.69; 0.22)] (Figure S4).

When only hormone-naïve TM studies were included, the absence of body fat gain remained up to one year of therapy compared to baseline [SMD: -0.20, 95% CI (-0.49; 0.08)], and a reduction was observed between one and two years [SMD: -0.41, 95% CI (-0.67; -0.15)] and after three years [SMD: -0.42, 95% CI (-0.78; -0.06)] (Figure S5).

Regarding puberty suppression, no temporal comparisons were possible. An increase in body fat mass was observed (SMD: 0.25, 95% CI [0.03; 0.46]) in TW who underwent puberty suppression, (Figure S6). Conversely, in TM who underwent puberty suppression, no differences in body fat mass were observed (SMD: 0.36, 95% CI [-0.05; 0.78]) (Figure S6).

#### Lean Mass

Of the 26 studies reporting this outcome for this comparison, 19 were included in the meta-analysis. Five studies were excluded because they either lacked extractable data (due to a graphical format) or did not report the duration of hormone therapy.

First, we conducted the analysis with all studies, regardless of whether the individuals were naïve or had undergone puberty suppression.

In TW, a decrease in lean mass was observed after one year of cross-sex hormone therapy compared to baseline [SMD: -0.22, 95% CI (-0.37; -0.08)] (Figure S7). No difference between two years and three years were observed when compared to baseline [SMD: -0.32, 95% CI (-0.76; 0.13)] (Figure S7).

When only studies with hormone-naïve TW were included, it was only possible to make a comparison after 1 year of therapy compared to baseline. A decrease in lean mass was observed [SMD: -0.26, 95% CI (-0.42; -0.09)] (Figure S8).

In TM, an increase in body lean mass was observed after one year of cross-sex hormone therapy compared to baseline [SMD: 0.51, 95% CI (0.39; 0.64)] (Figure S9). The same result was observed between one and two years [SMD: 0.39, 95% CI (0.24; 0.53)], and no differences were observed after three years when compared with baseline [SMD: 0.41, 95% CI (-0.09; 0.91)] (Figure S9).

When only hormone-naïve TM studies were included, an increase in body lean mass was observed after one year of hormone therapy compared to baseline [SMD: 0.55, 95% CI (0.39; 0.71)] (Figure S10). The same result was observed between one and two years [SMD: 0.44, 95% CI (0.27; 0.61)], and after three years [SMD: 0.57, 95% CI (0.21; 0.93)] (Figure S10).

Regarding puberty suppression, no temporal comparisons were possible. In both TW and TM who underwent puberty suppression, no significant differences were observed [SMD: 0.10, 95% CI (-0.44; 0.65) and SMD: -0.25, 95% CI (-0.62; 0.12)], from TW and TM, respectively) (Figure S11).

#### Upper-body strength

All 11 studies reporting this outcome for this comparison were included in the meta-analysis.

First, we conducted the analysis with all studies reporting this outcome, regardless of whether the individuals were naïve for hormone therapy. It was not possible to conduct analysis for puberty suppression.

In TW, a decrease in upper-body strength was observed after one year of cross-sex hormone therapy compared to baseline [SMD: -0.34, 95% CI (-0.54; -0.14)] (Figure S12). No difference between two years and three years were observed when compared to baseline [SMD: -0.22, 95% CI (-1.13; 0.70)] (Figure S12).

When only studies with hormone-naïve TW were included, a decrease in upper-body strength was observed after one year of cross-sex hormone therapy compared to baseline [SMD: -0.35, 95% CI (-0.56;-0.14)] (Figure S13). The same result was observed after follow-ups between two and three years [SMD: -0.67, 95% CI (-1.11; -0.22)] (Figure S13).

In TM, an increase in upper-body strength was observed after one year of cross-sex hormone therapy compared to baseline [SMD: 0.73, 95% CI (0.63; 0.83)] (Figure S14). A similar result was observed after follow-ups between one and two years [SMD: 0.91, 95% CI (0.35; 1.46)] and after three years when compared with baseline [SMD: 1.10, 95% CI (0.34; 1.87)] (Figure S14).

When only hormone-naïve TM studies were included, an increase in upper-body strength was observed after one year of cross-sex hormone therapy compared to baseline [SMD: 0.76, 95% CI (0.65; 0.86)] (Figure S15). Increases in strength were also observed after follow-ups between one and two years [SMD: 0.91, 95% CI (0.35; 1.46)], and no differences were observed after three years when compared with baseline [SMD: 1.03, 95% CI (0.00; 2.06)] (Figure S15).

#### Lower-body strength

The three studies reporting this outcome were included in the meta-analysis.

Analyses of lower-body strength were constrained by the limited number of available studies, precluding both temporal comparisons and assessments of puberty suppression - all studies included involved exclusively treatment-naïve patients. Cross-sex hormone therapy duration in TW and TM was 1 year in one study, ∼2.5 years in the other and one did not reported duration. No significant differences were observed in TW when compared to baseline (i.e., before cross-sex hormone therapy) [SMD: -0.17, 95% CI [-0.58; 0.25]) (Figure S16). An increase in lower-body strength was observed in TM when compared to baseline [SMD: 0.48, 95% CI (0.28; 0.69)] (Figure S16).

#### VO_2_ peak

Only one study reported VO□peak for these comparisons; therefore, a meta-analysis for this outcome was not performed.

### Risk of bias

Quality scores ranged from 10 to 18, with most studies rated as high (66.7 %; N = 10) or fair (20.0 %; N = 3), and 13.3 % as low quality (N = 2) for cross-sectional studies. Using AXIS, the most common weaknesses identified in the articles were the lack of justification of the sample size (N = 13) and the use of a convenience sample (N = 14).

Among cohort and quasi-experimental studies, 65.6% exhibited moderate risk of bias, with limitations concerning confounding (15/32 studies, 46.8%, moderate risk) and selective reporting (22/32 studies, 68.8%, moderate risk). Bias in participant selection, intervention classification and deviations from intended interventions were predominantly low risk (24/32, 75%; 25/32, 78.1%; 30/32, 93.8%, respectively).

All RCTs were classified as some concerns. Methodological concerns primarily stemmed from selection of the reported result (3/3 studies, 100%, some concerns); deviation from intended interventions (3/3 studies, 100%, some concerns); and randomization process (2/3 studies, 66.6%, some concerns).

A detailed assessment of the risk of bias for individual studies is provided in the supplementary material (Table S3 and S4, Figure S17).

## Discussion

### Statement of principal findings

In addition to evaluating physical fitness and body composition differences between transgender and cisgender individuals, this systematic review and meta-analysis also aimed to examine the influence of GAHT duration on the outcomes, assess variations between TM and TW in response to therapy, compare naïve vs. non-naïve individuals, and explore the impact of puberty suppression on body composition and strength. Furthermore, the review evaluated the methodological rigor and risk of bias in existing studies to provide a comprehensive understanding of the evidence quality and practical implications for the findings.

The meta-analysis of 20 studies revealed that TW exhibited significantly higher fat mass than CM (SMD: 0.85), but similar to CW. In respect of lean mass, meta-analysis of 21 studies revealed that TW showed higher values than CW (SMD: 0.98), but lower than CM (SMD: -1.17). Of relevance, the meta-analysis of 12 studies for upper-body strength and four for lower-body strength revealed that both were not significantly different between TW and CW (SMD: 0.41 and 0.05, respectively), but were markedly reduced in TW compared to CM (SMD: -1.79 and -1.99, respectively). Longitudinal hormone therapy (1–3 years) led to progressive increases in TW’s fat mass (SMD: 0.44 to 0.70) and declines in both lean mass (SMD: -0.22 to -0.32) and upper-body strength (SMD: -0.34 to -0.67). VO□peak in TW did not differ from CW, but was lower than CM (SMD: -1.61).

The meta-analysis comparing TM and CM revealed significant differences, with TM showing intermediate body composition and strength metrics between CW and CM, even after an average therapy duration of 4 years.

### Practical implications

This review shows that, despite TW exhibiting higher absolute lean mass compared to CW, no significant differences in upper- or lower-body strength were observed between the two groups after 1–3 years of hormone therapy. This finding challenges the assumption that potential residual lean mass inherently translates to functional strength advantages in this population. For example, TW’s upper-(SMD: 0.41, 95% CI [-0.10; 0.92]) and lower-body strength (SMD: 0.05, 95% CI [-0.74; 0.83]) showed negligible divergence from CW, even as total body lean mass declined modestly in response to GAHT (SMD: -0.22 to -0.32). These results align with evidence [4] showing that, while TW retain higher absolute lean mass, body composition-adjusted strength metrics (e.g., relative strength per kg lean mass) converge with CW over time. This suggests that lean mass alone is an incomplete proxy for TW’s athletic performance, as neuromuscular efficiency, training history, and fat distribution may play compensatory roles. In fact, when height-normalized, TW and CW appear to show comparable appendicular lean mass/height² or lean mass/height² [15, 66]. Importantly, a greater absolute lean mass not accompanied by increased functionality may actually impair performance, especially in weight-sensitive sports (e.g. cycling and climbing), which not surprisingly have the lightest athletes[67].

Furthermore, sport performance extends beyond physiology to encompass social, psychological, and cultural dimensions (e.g., stigmatization, discrimination, access to sports opportunities, self-concept, self-esteem etc), which altogether may influence athletic engagement and achievement among transgender individuals. Indeed, evidence indicates that this population faces an elevated risk of adverse mental health outcomes [68], likely due to systemic stigma and discrimination across various contexts [69], including sports [70, 71]. While the extent to which potential “muscle memory” (i.e., long-lasting physiological effects of prior testosterone exposure) may counteract the influence of these psychosocial factors on athletic performance remains unestablished, the assumption of inherent competitive advantages for TW over CW does not appear to be robustly supported by existing evidence.

In fact, the absence of strength disparities between TW and CW found in the current review contradicts narratives framing male puberty as conferring irreversible athletic advantages despite GAHT. In a narrative review, Lundberg et al. (2024) [3] argue that male developmental traits (e.g., height, skeletal proportions) inherently disrupt fairness, yet the lack of measurable strength differences in the present systematic review suggests such claims may overemphasize structural factors while underestimating the impact of GAHT. For instance, TW’s VO□peak, when adjusted for weight, aligns with CW [4], further supporting parity in endurance capabilities. Furthermore, TW’s pre-therapy advantages in push-ups and sit-ups disappeared after 2 years of feminizing hormones among 46 individuals who started GAHT while in the United States Air Force [38]. These findings are corroborated by the current meta-analysis, endorsing nuanced, sport-specific policies rather than blanket bans.

### Limitations of the available evidence and the review

This systematic review aligns with previous ones [1, 4] in highlighting critical research limitations. This includes the typically short study durations (<3 years) and a lack of data on elite athletes. Additionally, the potential conflation of trained and untrained individuals complicates extrapolation. The available evidence remains limited for specific outcomes (e.g., lower-body strength and VO□peak), particularly regarding RCTs examining the effects of GAHT on physical fitness and body composition (n = 3), as well as studies assessing the impact of puberty suppression (n = 6). Another literature weakness is the inconsistent reporting and adjustment for confounders, as few studies controlled for training history, diet, baseline fitness, physical activity and body composition or previous hormone therapy, potentially hindering the isolated effects of GAHT. Finally, there is very little literature involving transgender athletes of any age, across all sport settings, and at any competitive level. Therefore, future studies must prioritize transgender athletes, assess sport-specific performance metrics, and evaluate long-term (e.g., >5 years) physiological and psychological changes, controlling for puberty suppression whenever possible.

The limitations of this review are related to the identified gaps in literature and include: the reliance on short-term assessments, limiting conclusions about the effects of GAHT on targeted outcomes in the long run; the heterogeneity of the studies assessed, variability in hormone regimens (e.g., types/doses of antiandrogens, estrogens), measurement methods (e.g., DEXA vs. MRI for body composition), and control groups (e.g., inconsistent physical activity tracking); the reliance on lean mass and strength as proxies for performance, rather than sport-specific outcomes (e.g., race times, power output), which limits practical relevance to real-world sport scenario; underrepresentation of puberty-suppressed cohorts, hampering the meta-analytic comparison between suppressed vs. non-suppressed individuals; and the inclusion of studies with risk of bias and with cross-sectional or retrospective designs.

## Conclusions and perspectives

This systematic review and meta-analysis shows that, while TW exhibited higher absolute lean mass compared to CW, no significant differences in physical fitness metrics (i.e., upper-body strength, lower-body strength and VO□peak) were observed after 1–3 years of therapy. Although the current data do not justify blanket bans, critical gaps in literature were found, notably the underrepresentation of transgender athletes who may retain more “muscle memory”. Ideally, to resolve speculation, future long-term, longitudinal studies should prioritize performance-specific metrics in transgender athletes. However, one should be aware of the scarce number of transgender athletes, particularly in the elite sport, which complicates the feasibility of conducting powered studies involving high-performance transgender athletes within specific sport disciplines. In light of this context of imperfect evidence and despite the methodological challenges, continued research into physiological as well as psychosocial trajectories among transgender athletes with diverse demographics and clinical characteristics remains essential for developing equitable frameworks that balance justice, inclusion, and scientific rigor. Policies should remain dynamic, guided by evolving evidence and ethical imperatives, whereas acknowledging that fairness and non-discrimination are interdependent objectives necessitating nuanced, context-sensitive strategies.

## Data Availability

All data produced in the present study are available upon reasonable request to the authors.

## Funding

Authors were supported by São Paulo Research Foundation (FAPESP) and Coordination for the Improvement of Higher Education Personnel (CAPES).

## Competing interests

All authors confirm that there are no conflicts of interest associated with this work.

## Contributorship

SMS and DCR searched studies in the databases, ran statistical analysis, elaborated results and drafted the manuscript. SMS, BCM, DCR, IL, ARC and JFVMPH extracted data. BCM and IL evaluated risk of bias, edited and revised the manuscript. BG conceived the study design and drafted the manuscript. HR conceived the study design and revised the manuscript. All authors contributed to critical revision of the report for important intellectual content.

## Acknowledgements

None to declare.

## Data sharing statement

All data produced in the present study are available upon reasonable request to the authors.

## Ethics approval

Not applicable.

## Patient and public involvement

Patients and/or the public were not involved in the design, or conduct, or reporting, or dissemination plans of this research.

## Notes

### Competing Interest Statement

The authors have declared no competing interest.

### Funding Statement

Authors were supported by Sao Paulo Research Foundation (FAPESP) and Coordination for the Improvement of Higher Education Personnel (CAPES).

