## supplementary material for "Physical Fitness and Body Composition in Transgender vs. Cisgender Individuals: A Systematic Review and Meta-Analysis"

**Supplementary Materials**

Table S1. Search strategy

| Descriptors | Terms |
| --- | --- |
| Population #1 | transgender OR transexual OR “transgender person” OR transsexualism OR transgenderism OR “transgender people” OR “gender reassignment procedure” OR “gender reassignment surgery” OR “gender change procedure” OR “gender-affirming treatment” OR transwoman OR transmen OR “gender affirming hormone therapy” OR transsexual OR “cross-sex hormone therapy” OR “trans people” |
| Outcome #2 | "muscle tonus" OR "treadmill tests" OR "cardiopulmonary exercise test" OR "exercise test" OR "psychomotor performance" OR "physical endurance" OR "physical functional performance" OR "athletic performance" OR "cardiorespiratory fitness" OR "muscle strength" OR "physical fitness" OR “muscle strength dynamometer” OR “endurance shuttle walk test” OR “incremental shuttle walk test” OR “6-minute walk test” OR “arm ergometry test” OR “bicycle ergometry test” OR “physical fitness testing” OR “eurofit test” OR “muscle power” OR “1RM” OR “one repetition maximum” OR “vertical jump” OR “wingate anaerobic” OR “isometric mid-thigh pull” OR “countermovement jump” OR CMJ OR “physical test*” OR “step test” OR “VO2” OR “VE/VO2” OR “respiratory exchange ratio” OR “grip strength” OR handgrip OR dynamometer OR “explosive strength” OR “sit-up*” OR “1.5 mile run time” OR “push up*” OR “jump height” OR “lower body anaerobic power” OR “anaerobic power” OR “sit and reach test” OR “range of motion” OR “cardiorespiratory fitness” OR “cardiorespiratory endurance” OR “muscular fitness” OR “musculoskeletal fitness” OR “muscular strength” OR “muscular endurance” OR “flexibility test*” OR sprint OR agility OR “change-of-direction” OR “force–velocity test” OR “squat jump” OR “speed” OR “velocity” OR “upper body strength” OR “lower body strength” OR “body composition” OR “body mass” OR “body weight” BMI OR “body fat” OR “lean body mass” OR “cross-sectional area” OR “muscular area” OR "skeletal muscle" OR "muscle mass" OR "lean mass" OR "appendicular lean mass" OR “abdominal visceral fat” OR “abdominal subcutaneous fat” |
| Combination | #1 AND #2 |

**Table S2. Therapy characteristics.**

| **Author (date)** | **Therapy** | **Type** | **Duration**  **(years)** | **Other intervention** | **Hormone**  **naïve at baseline** | **Adverse event** | **Dropout** |
| --- | --- | --- | --- | --- | --- | --- | --- |
| Alvares et al. (2025) | GAHT | NR | ~7 | - | - | - | - |
| Saitong et al. (2025) | GAHT | EV  17ß estradiol  CPA | >1 | GAS | - | - | - |
| Amador et al (2024) | GAHT | EV  CPA | ~3 | None | - | - | - |
| Ceolin et al (2024) | GAHT | NR | NR | GAS | - | - | - |
| Ciancia et al (2024) | GnRHa  GAHT | Triptorelin  TW: EV  TM: Testosterone | ~3 | Puberty suppression | NR | NR | NR |
| Hamilton et al (2024) | GAHT | NR | TW  6±4  TM  4±5 | - | - | - | - |
| Pei et al (2024) | GAHT  Antiandrogen | EV  Estradiol gel  CPA | 1-3 | None | Yes | NR | NR |
| Boogers et al (2023) | GnRHa  GAHT | Triptorelin  TW: Estradiol  TM: Testosterone | 3 | Puberty suppression | Yes | NR | TW: 43  TM: 74 |
| Chiccarelli et al (2023) | GAHT | NR | NR | NR | NR | NR | NR |
| Tominaga et al (2023) | GAHT | Testosterone  LD  HD | 3 months to 10 y | NR | Yes | None | NR |
| Yaish et al (2023) | GAHT  Antiandrogen | Estradiol  CPA | 6 months | NR | Yes | None | None |
| Yamada et al (2023) | GAHT | Testosterone | 1 | GAS | NR | NR | NR |
| Alvares et al (2022) | GAHT | Testosterone | ~2 | None | - | - | - |
| Andrade et al (2022) | GAHT  Antiandrogen | Estrogen  CPA | 14±4 | None | - | - | - |
| Klaver et al (2022) | GAHT  Antiandrogen | TW: EV  Transdermal Estradiol  CPA  TM: Testosterone  TU  TE | 1 | None | Yes | NR | None |
| Bretherton et al (2021) | GAHT | TW: Estradiol  TM: Testosterone | ~4 | None | - | - | - |
| Gava et al (2021) | GAHT | TU+PL  TU+DT | ~1 | GAS | No. * A washout period  of 12 weeks was required | None | 2 |
| Navabi et al (2021) | GnRHa | Leu | NR | Puberty suppression | Yes | NR | NR |
| Nokoff et al (2021) | GnRHa | NR | TM  21±20 months  TW  11± 7 months | None | - | - | - |
| Yun et al (2021) | GAHT  Antiandrogen | EV  CPA  Spironolactone | 6 months | None | Yes | NR | NR |
| Gava et al (2020) | GAHT  Antiandrogen | CPA+E  Leu+ E | 5 | NR | Yes | NR | NR |
| Jenkins et al (2020) | GnRHa  GAHT  Antiandrogen | NR | 2 | - | - | - | - |
| Nokoff et al (2020) | GAHT | TW: Estradiol  TM: Testosterone | ~1 | Puberty suppression  GnRHa in the past  GAS | - | - | - |
| Roberts et al (2020) | GAHT  Antiandrogen | TW: Estradiol  Transdermal estradiol  Estradiol cypionate  EV  Spironolactone  Finasteride  TM: testosterone cypionate  TE | ~2.5 | None | Yes | NR | None |
| Shadid et al (2020) | GAHT  Antiandrogen | TW: EV  Estradiol  Transdermal estradiol  CPA  TM: Testosterone IM | 1 | NR | Yes | NR | NR |
| Velzen et al (2020) | GAHT  Antiandrogen | TM: Testosterone IM  TU  TW: Estradiol  CPA | 2 | NR | Yes | NR | NR |
| Chrisostomo et al (2019) | GAHT  Antiandrogen | EV  CPA  Spironolactone | 3 | None | - | - | - |
| Scharff et al (2019) | GAHT  GAHT  Antiandrogen | TW: EV  Estradiol  Transdermal Estrogens  CPA  TM: Testosterone  TE  TU | 1 | NR | Yes | NR | NR |
| Wiik et al (2019) | GnRHa  GAHT | TW: Estradiol  TM: TU IM | 1 | NR | Yes | NR | NR |
| Auer et al (2018) | GAHT  Antiandrogen | TW: Estradiol  CPA  TM: Testosterone | 1 | None | Yes | NR | NR |
| Fighera et al (2018) | GAHT | EV  Spironolactone | 3 months  3 years | GAS | No | NR | TW: 67.5% |
| Gava et al (2018) | GAHT | TU  TE | 5 | GAS | Yes | NR | NR |
| Klaver et al (2018) | GnRHa  GAHT | NR | NR | None | Yes | NR | TW  start of GnRHa: 12%  start of CHT: 36%  22y: 64%  TM  start of GnRHa: 11%  start of CHT: 45%  22y: 65% |
| Tack et al (2018) | GAHT | Lynestrenol  CPA | ~1 | Puberty suppression | NR | NR | None |
| Hannema et al (2017) | GnRHa  GAHT | Triptorelin  Estradiol | 3 | GAS | No | None | None |
| Klaver et al (2017) | GAHT  Antiandrogen | TW: Estradiol  CPA  TM:TE  TU | 1 | NR | Yes | NR | None |
| Auer et al (2016) | GAHT  Antiandrogen | TW: Estradiol  CPA  TM: Testosterone | 1 | NR | TW: Yes  TM: 6 had already received | NR | NR |
| Caenegem et al (2015)a | GAHT | EV  transdermal 17-β estradiol  CPA | 1-2 | None | NR | NR | 20 |
| Caenegem et al (2015)b | GAHT | TU | 1 | None | NR | NR | None |
| Klink et al (2015) | GnRHa  GAHT | Triptorelin  TW: Estradiol  TM: TE | Start of GnRHa  11-18 years old  Start of CHT  16-19 years old | Puberty suppression | Yes | NR | NR |
| Pelusi et al (2014) | GAHT | Testoviron Depot  Testosterone  TU | ~1 | None | Yes | None | NR |
| Vilas et al (2014) | GAHT  Antiandrogen | TW: EV  CPA  TM: TU | 20±31 months | None | No | NR | None |
| Wierckx et al (2014) | GAHT  Antiandrogen | TW: EV  CA  CPA  transdermal 17‐β estradiol  TM: TU IM | 1 | None | Yes | TM  muscle/joint aches, mild blood pressure, reduced fasting insulin, androgenetic alopecia.  TW  depression, elevated prolactin, galactorrhea, Transient liver enzyme elevations, hypertension, fasting insulin increased, skin irritation. | NR |
| Caenegem et al (2013) | GAHT | NR | NR | GAS | - | - | - |
| Caenegem et al (2012) | GAHT | Testosterone | 10 years old  (3–28) | None | - | - | - |
| Mueller et al (2010)a | GAHT | TU | 2 | None | Yes | None | None |
| Mueller et al (2010)b | GnRHa  GAHT | Goserelin acetate  estradiol-17 β valerate | 2 | None | Yes | None | None |
| Lapauw et al (2008) | GAHT | NR | 8 years old  (4, 20) | GAS | - | - | - |
| Meriggiola et al (2008) | GAHT | Testosterone | ~1 | None | No | None | None |
| Elbers et al (1999) | GAHT  Antiandrogen | TW: Ethinyl estradiol  CPA  TM: Testosterone | 1 | None | NR | - | NR |
| Elbers et al (1997) | GAHT | Testosterone | 3–4 | GAS | NR | - | NR |

Legend: GAHT: gender-affirming hormone therapy; GAS - gender-affirming surgery; GnRH :gonadotropin releasing hormone; GnRHa :gonadotropin releasing hormone antagonist; LD: low dose of testosterone; HD: high dose of testosterone; EV: estradiol valerate; TE: testosterone enanthate; TU: testosterone undecanoate; CPA: cyproterone acetate; E: estradiol; Leu: leuprolide acetate; DT: dutasteride; PL: placebo; SLE: estradiol only protocol; CO: combined oral; IM: intramuscular

**Table S3. Quality assessment of cross-sectional studies using AXIS**

| **Basic Information** | **Introduction** | **Methods** | | | | | | | | | | **Results** | | | | | **Discussion** | | **Other** | | **Total score** | **Classification** |
| --- | --- | --- | --- | --- | --- | --- | --- | --- | --- | --- | --- | --- | --- | --- | --- | --- | --- | --- | --- | --- | --- | --- |
| **Author/Year** | **Q1** | **Q2** | **Q3** | **Q4** | **Q5** | **Q6** | **Q7** | **Q8** | **Q9** | **Q10** | **Q11** | **Q12** | **Q13** | **Q14** | **Q15** | **Q16** | **Q17** | **Q18** | **Q19** | **Q20** |  |  |
| Alvares et al 2022 | 1 | 0 | 0 | 1 | 0 | 1 | 1 | 1 | 1 | 1 | 1 | 0 | 0 | 1 | 1 | 0 | 1 | 1 | 1 | 1 | 14 | High |
| Amador et al 2024 | 1 | 1 | 0 | 1 | 0 | 1 | 1 | 1 | 1 | 1 | 1 | 0 | 0 | 1 | 1 | 1 | 1 | 0 | 1 | 1 | 15 | High |
| Andrade et al 2022 | 0 | 0 | 0 | 0 | 0 | 0 | 1 | 0 | 1 | 1 | 1 | 0 | 0 | 1 | 1 | 0 | 1 | 1 | 1 | 1 | 10 | Low |
| Bretherton et al 2021 | 1 | 0 | 0 | 1 | 0 | 1 | 1 | 1 | 1 | 1 | 1 | 0 | 0 | 1 | 0 | 0 | 0 | 1 | 1 | 1 | 12 | Fair |
| Caenegem et al 2012 | 1 | 0 | 0 | 1 | 0 | 1 | 1 | 1 | 1 | 1 | 1 | 1 | 0 | 1 | 1 | 1 | 1 | 1 | 0 | 1 | 15 | High |
| Caenegem et al 2013 | 0 | 0 | 0 | 1 | 0 | 0 | 1 | 0 | 1 | 1 | 1 | 1 | 0 | 1 | 1 | 1 | 1 | 0 | 1 | 1 | 12 | Fair |
| Ceolin et al 2024 | 0 | 0 | 0 | 1 | 0 | 1 | 1 | 1 | 1 | 1 | 1 | 0 | 0 | 1 | 1 | 0 | 1 | 1 | 1 | 1 | 13 | Fair |
| Chrisostomo et al 2019 | 1 | 1 | 0 | 1 | 0 | 1 | 0 | 1 | 1 | 1 | 1 | 0 | 0 | 0 | 0 | 0 | 0 | 0 | 0 | 1 | 9 | Low |
| Hamilton et al 2024 | 1 | 1 | 0 | 1 | 0 | 0 | 1 | 1 | 1 | 1 | 1 | 0 | 0 | 1 | 1 | 1 | 0 | 1 | 1 | 1 | 14 | High |
| Jenkins et al 2020 | 1 | 1 | 1 | 1 | 0 | 1 | 1 | 1 | 1 | 1 | 1 | 0 | 0 | 1 | 0 | 1 | 1 | 1 | 0 | 1 | 15 | High |
| Lapauw et al 2008 | 1 | 1 | 0 | 1 | 0 | 1 | 1 | 1 | 1 | 1 | 1 | 1 | 0 | 1 | 1 | 1 | 1 | 1 | 1 | 1 | 17 | High |
| Nokoff et al 2020 | 1 | 1 | 0 | 1 | 0 | 1 | 0 | 1 | 1 | 1 | 1 | 1 | 0 | 1 | 1 | 0 | 1 | 1 | 1 | 1 | 15 | High |
| Nokoff et al 2021 | 1 | 1 | 0 | 1 | 0 | 1 | 0 | 1 | 1 | 1 | 1 | 1 | 0 | 1 | 1 | 0 | 1 | 1 | 1 | 1 | 15 | High |
| Alvares et al (2025) | 1 | 1 | 0 | 1 | 0 | 1 | 1 | 1 | 1 | 1 | 1 | 1 | 1 | 1 | 1 | 1 | 1 | 1 | 0 | 1 | 17 | High |
| Saitong et al (2025) | 1 | 1 | 1 | 1 | 0 | 1 | 1 | 1 | 1 | 1 | 1 | 1 | 0 | 1 | 1 | 1 | 1 | 1 | 1 | 1 | 18 | High |
| Yamada et al 2023 | 1 | 1 | 1 | 1 | 0 | 1 | 1 | 1 | 1 | 1 | 1 | 1 | 1 | 1 | 1 | 1 | 1 | 1 | 1 | 1 | 19 | High |

Legend: AXIS: Appraisal tool for cross-sectional studies.

**Table S4. Quality assessment of cohorts and quasi-experimental studies using ROBINS-I**

| **Study** | **Bias due to confounding** | **Bias in selection of participants into the study** | **Bias in classification of interventions** | **Bias due to deviations from intended interventions** | **Bias due to missing data** | **Bias in measurement of outcomes** | **Bias in selection of the reported result** | **Overall Bias** |
| --- | --- | --- | --- | --- | --- | --- | --- | --- |
| Auer et al., 2016 | Moderate | Low | Low | Low | Moderate | Moderate | Moderate | Moderate |
| Auer et al., 2018 | Moderate | Low | Low | Low | Moderate | Low | Moderate | Moderate |
| Boogers et al., 2023 | Low | Moderate | Low | Low | Moderate | Low | Moderate | Moderate |
| Caenegem et al., 2015a | Low | Moderate | Low | Low | Low | Moderate | Moderate | Moderate |
| Caenegem et al., 2015b | Low | Moderate | Low | Low | Low | Moderate | Moderate | Moderate |
| Chiccarelli et al., 2023 | Low | Serious | Moderate | Low | Moderate | Moderate | Moderate | Serious |
| Ciancia et al., 2024 | Low | Low | Low | Low | Low | Low | Moderate | Moderate |
| Elbers et al., 1997 | Moderate | Low | Low | Moderate | Low | Low | Moderate | Moderate |
| Elbers et al., 1999 | Moderate | Low | Low | Low | Low | Low | Moderate | Moderate |
| Fighera et al., 2018 | Moderate | Moderate | Serious | Low | Low | Low | Serious | Serious |
| Gava et al., 2018 | Moderate | Low | Moderate | Low | Low | Low | Moderate | Moderate |
| Gava et al., 2020 | Low | Low | Low | Low | Moderate | Low | Moderate | Moderate |
| Hannema et al., 2017 | Serious | Serious | Serious | Low | Serious | Low | Moderate | Serious |
| Klaver et al., 2017 | Moderate | Low | Low | Low | Moderate | Low | Moderate | Moderate |
| Klaver et al., 2018 | Moderate | Low | Low | Low | Low | Low | Moderate | Moderate |
| Klaver et al., 2022 | Low | Low | Low | Low | Low | Moderate | Moderate | Moderate |
| Klink et al., 2015 | Serious | Serious | Moderate | Low | Low | Low | Low | Serious |
| Mueller et al., 2010a | Low | Low | Low | Low | Low | Low | Low | Low |
| Mueller et al., 2010b | Low | Low | Low | Low | Low | Low | Low | Low |
| Navabi et al., 2021 | Moderate | Serious | Moderate | Low | Serious | Low | Moderate | Serious |
| Roberts et al., 2020 | Moderate | Low | Moderate | Low | Low | Moderate | Moderate | Moderate |
| Scharff et al., 2019 | Low | Low | Low | Low | Moderate | Moderate | Serious | Serious |
| Schadid et al., 2020 | Moderate | Low | Low | Low | Low | Low | Low | Moderate |
| Tack et al., 2018 | Moderate | Low | Low | Low | Moderate | Moderate | Low | Moderate |
| Tominaga et al., 2023 | Low | Low | Low | Moderate | Serious | Moderate | Serious | Serious |
| van Velzen et al., 2020 | Moderate | Low | Low | Low | Low | Low | Moderate | Moderate |
| Wierckx et al., 2014 | Moderate | Low | Low | Low | Moderate | Low | Moderate | Moderate |
| Wiik et al., 2019 | Moderate | Low | Low | Low | Low | Moderate | Serious | Serious |
| Yaish et al., 2023 | Low | Low | Low | Low | Low | Low | Moderate | Moderate |
| Yun et al., 2021 | Moderate | Low | Low | Low | Moderate | Moderate | Low | Moderate |
| Vilas et al., 2014 | Serious | Low | Low | Low | Low | Low | Moderate | Serious |
| Pei et al., 2024 | Moderate | Low | Low | Low | Moderate | Moderate | Moderate | Moderate |

Legend: ROBINS-I: Risk Of Bias In Non-randomized Studies - of Interventions.

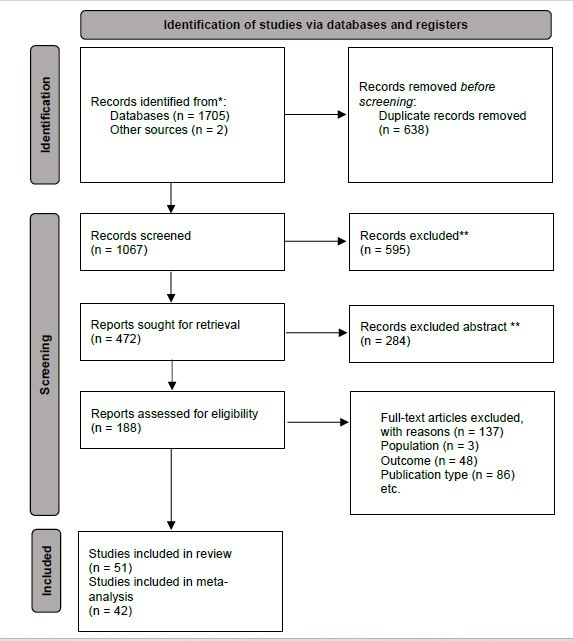

Figure S1. Flowchart showing the selection process for assessing studies.

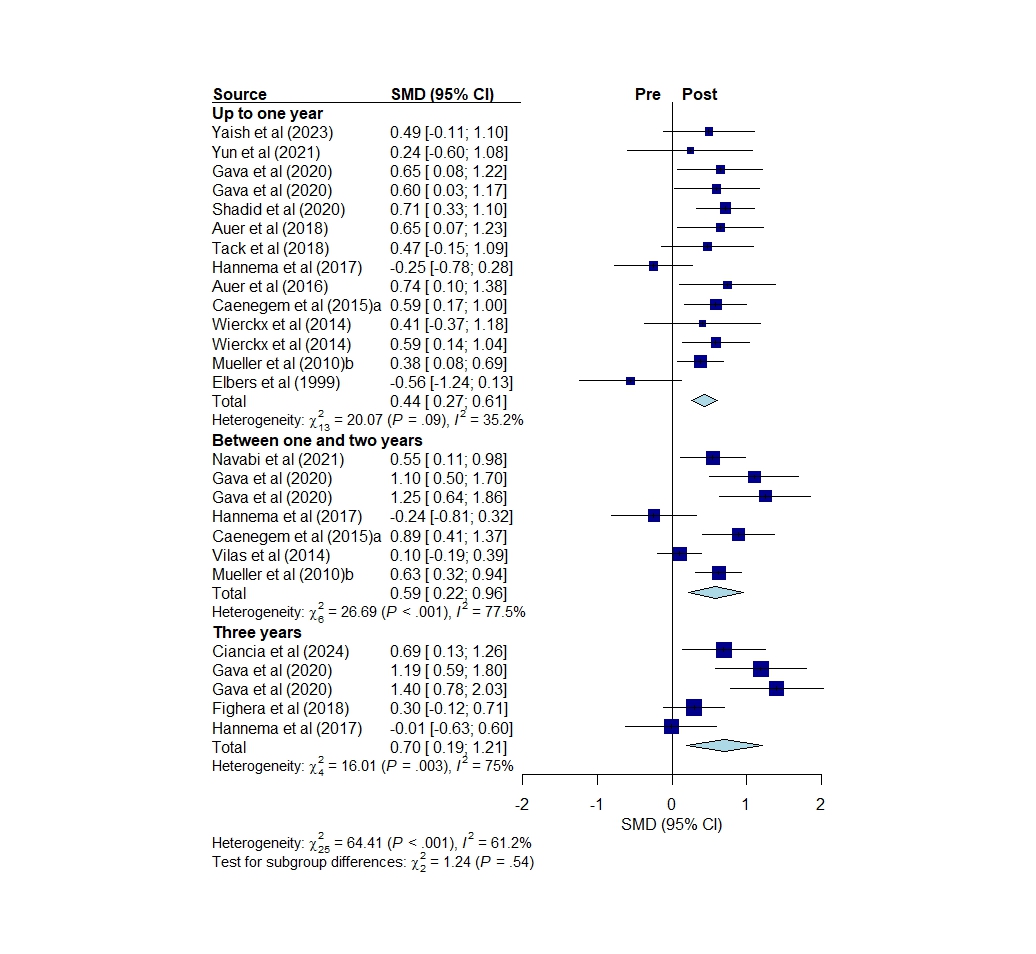

Figure S2. Forest plot: Fat mass pre vs. post hormone therapy in transgender women (all studies).

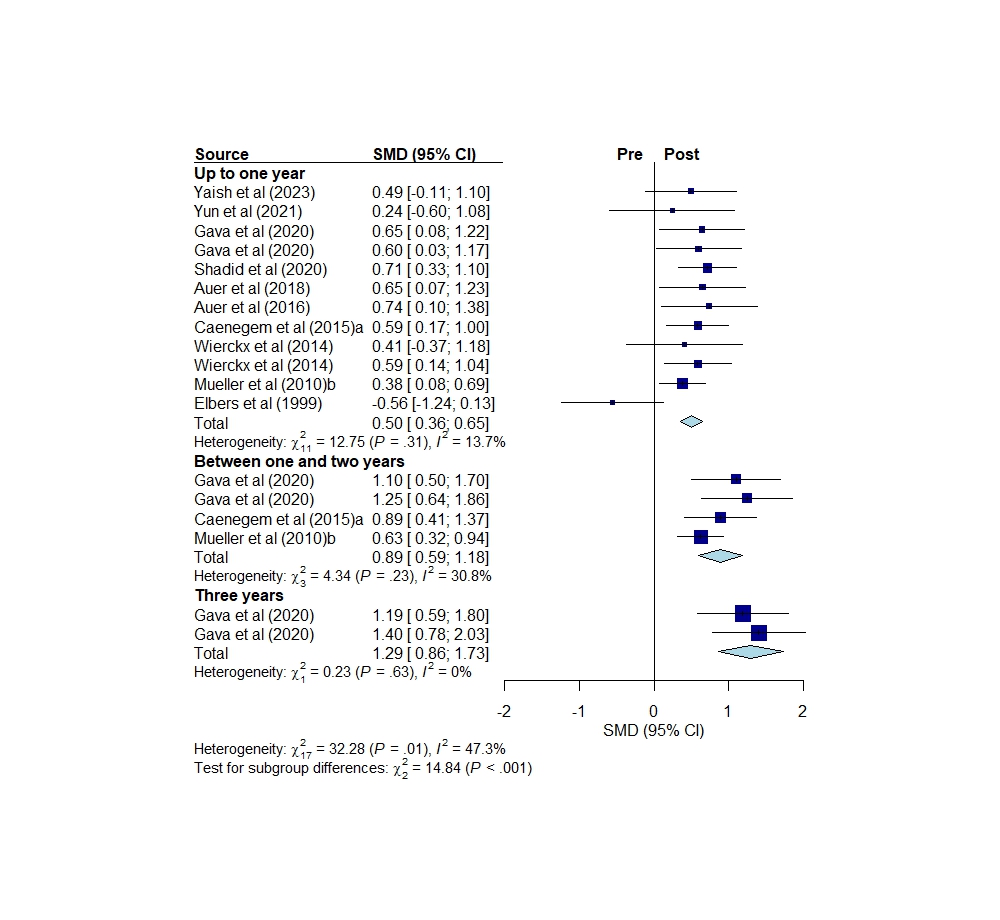

Figure S3. Forest plot: Fat mass pre vs. post hormone therapy in hormone-naïve transgender women.

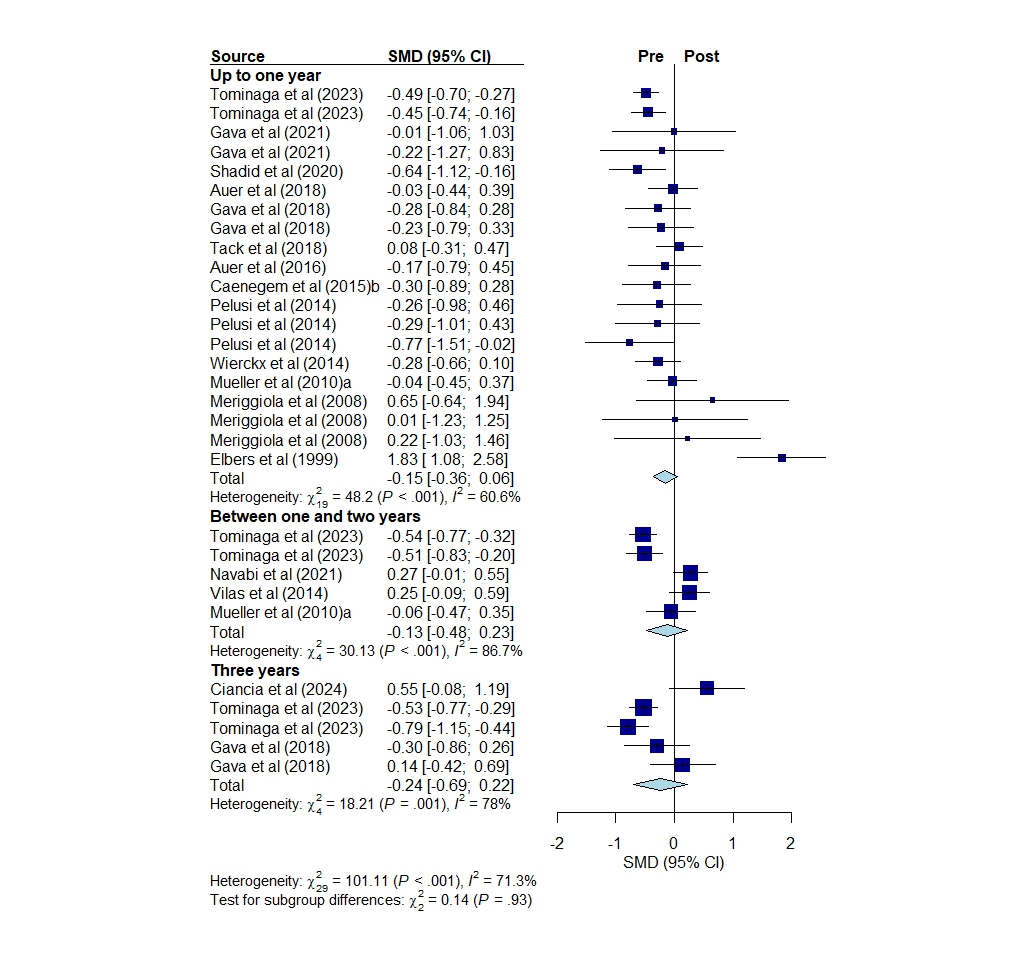

Figure S4. Forest plot: Fat mass pre vs. post hormone therapy in transgender men (all studies).

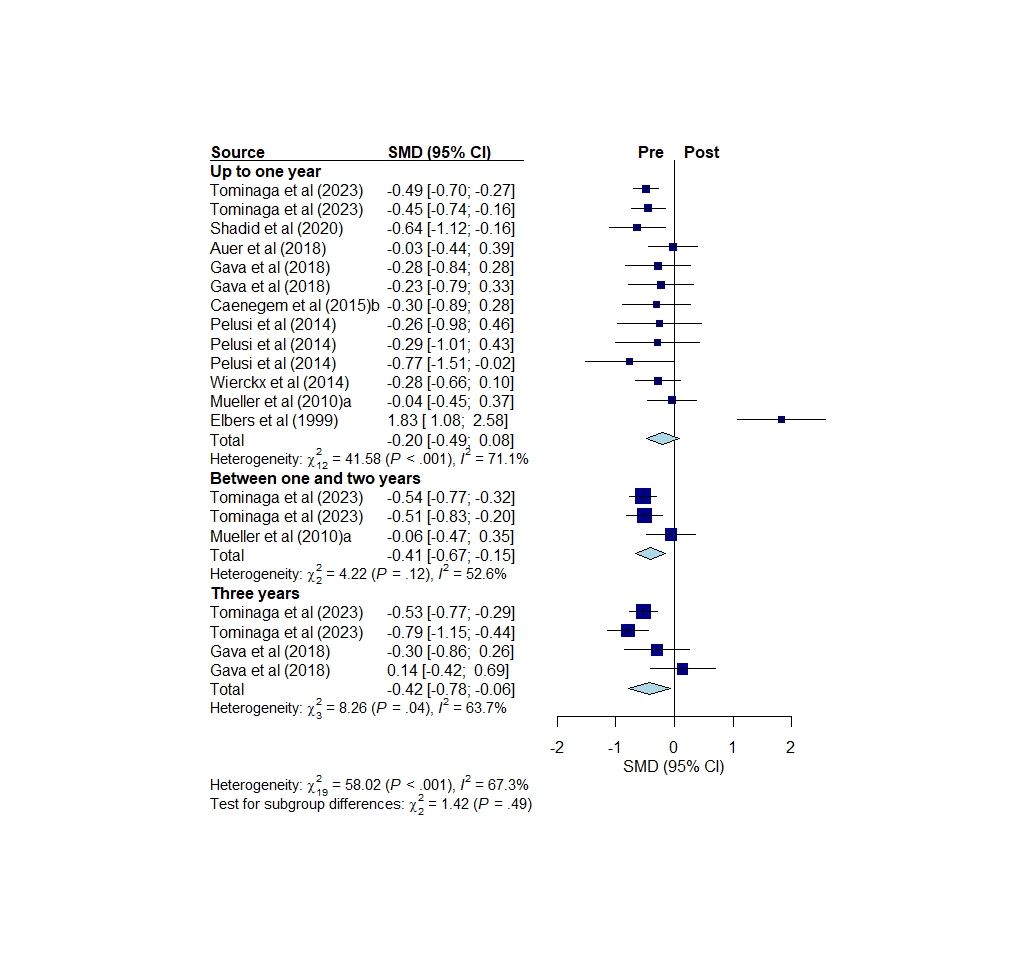

Figure S5. Forest plot: Fat mass pre vs. post hormone therapy in hormone-naïve transgender men.

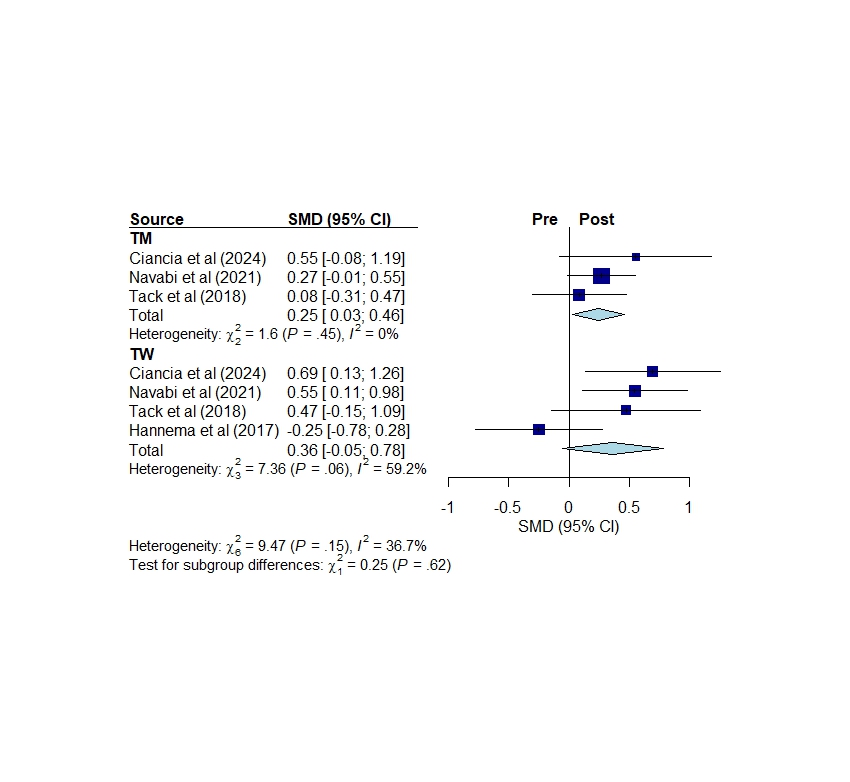
Figure S6. Forest plot: Fat mass in transgender men (TM) and women (TW) pre vs. post puberty suppression.

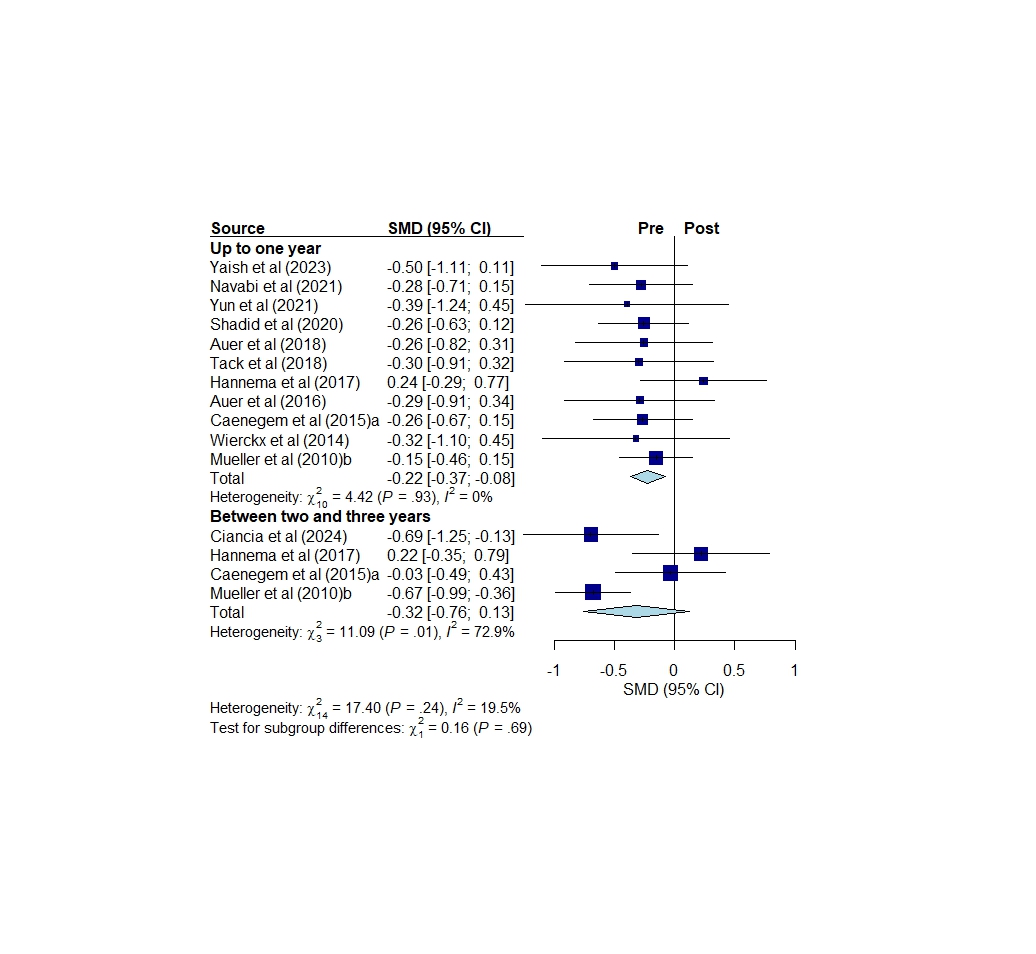

Figure S7. Forest plot: Lean mass pre vs. post hormone therapy in transgender women (all studies).

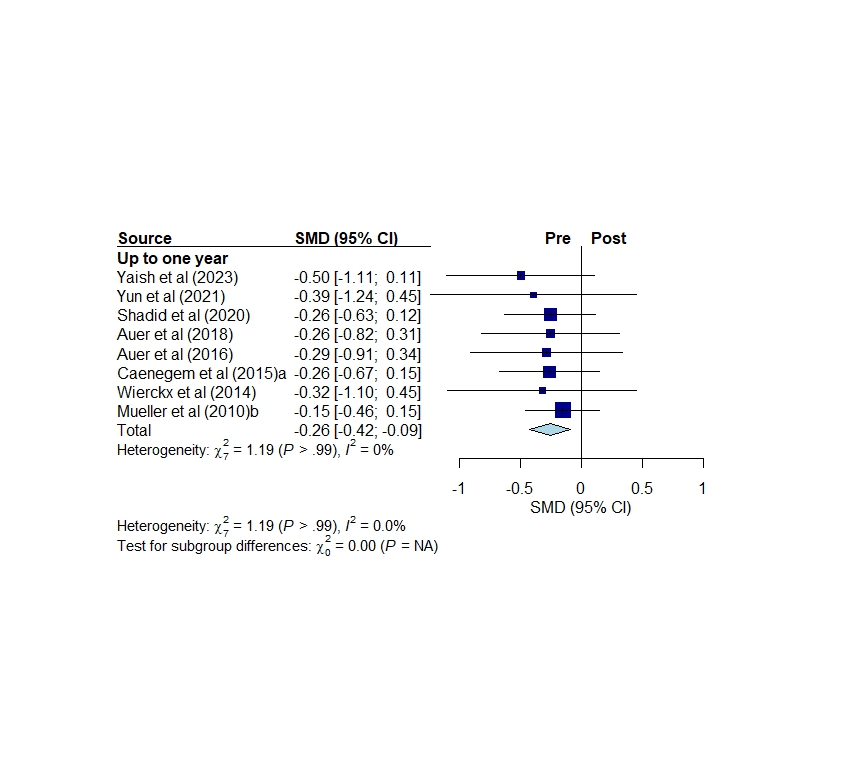

Figure S8. Forest plot: Lean mass pre vs. post hormone therapy in hormone-naïve transgender women.

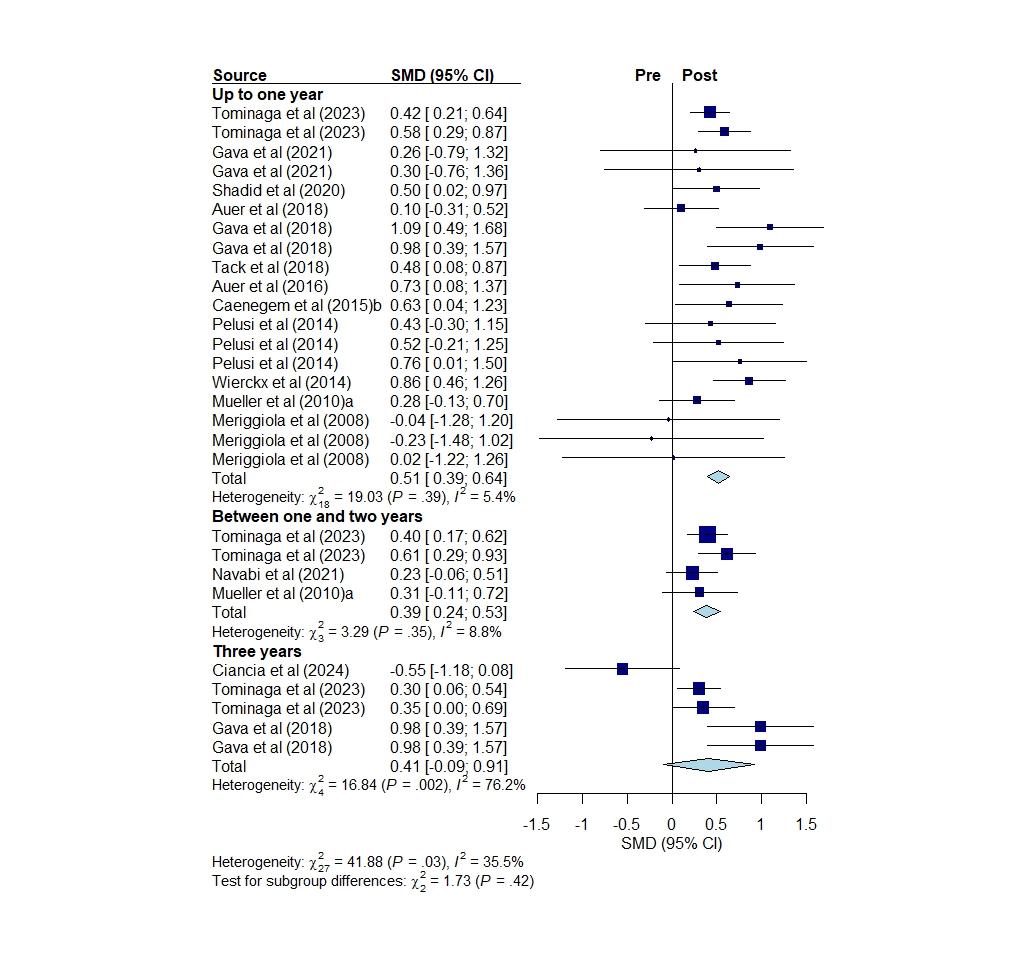

Figure S9. Forest plot: Lean mass pre vs. post hormone therapy in transgender men (all studies).

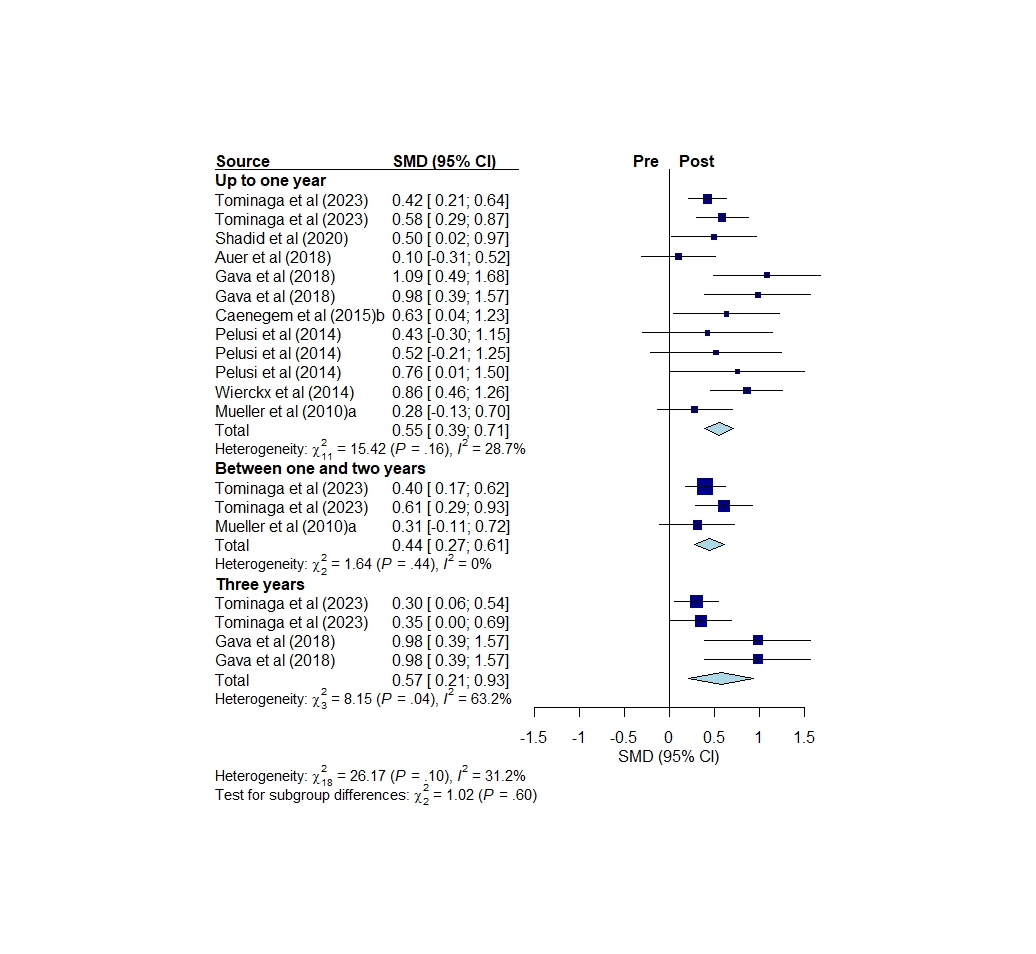

Figure S10. Forest plot: Lean mass pre vs. post hormone therapy in hormone-naïve transgender men.

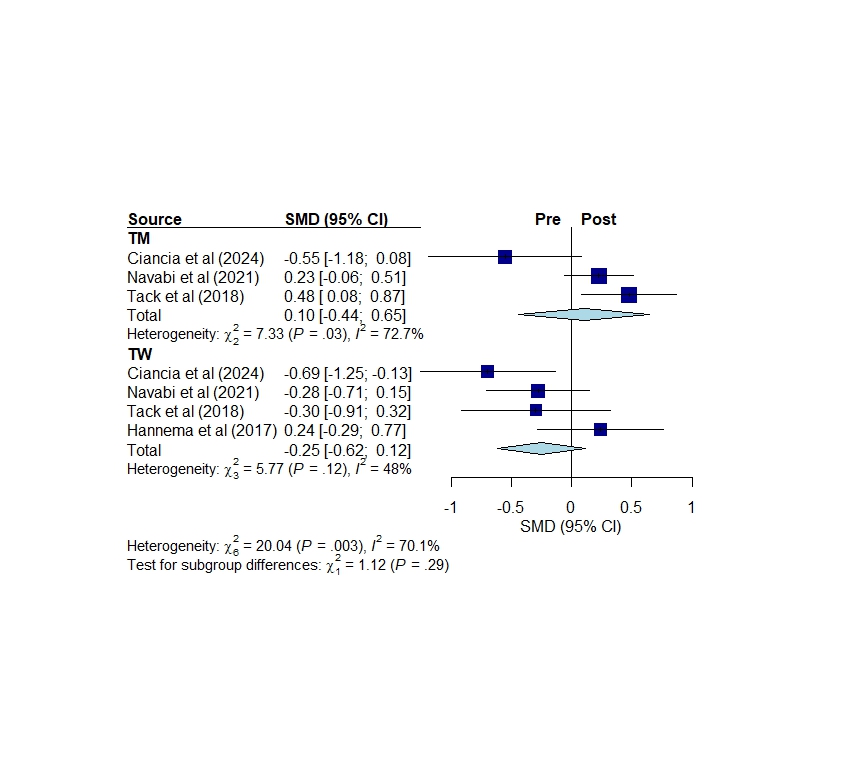

Figure S11. Forest plot: Lean mass in transgender men (TM) and women (TW) before vs. after puberty suppression.

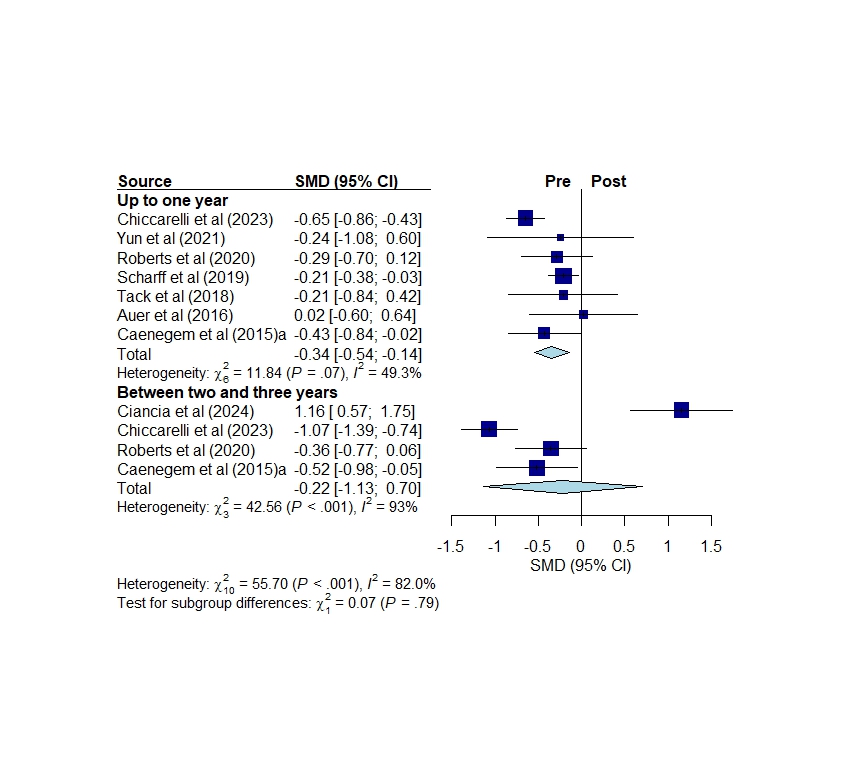

Figure S12. Forest plot: Upper-body strength pre vs. post hormone therapy in transgender women (all studies).

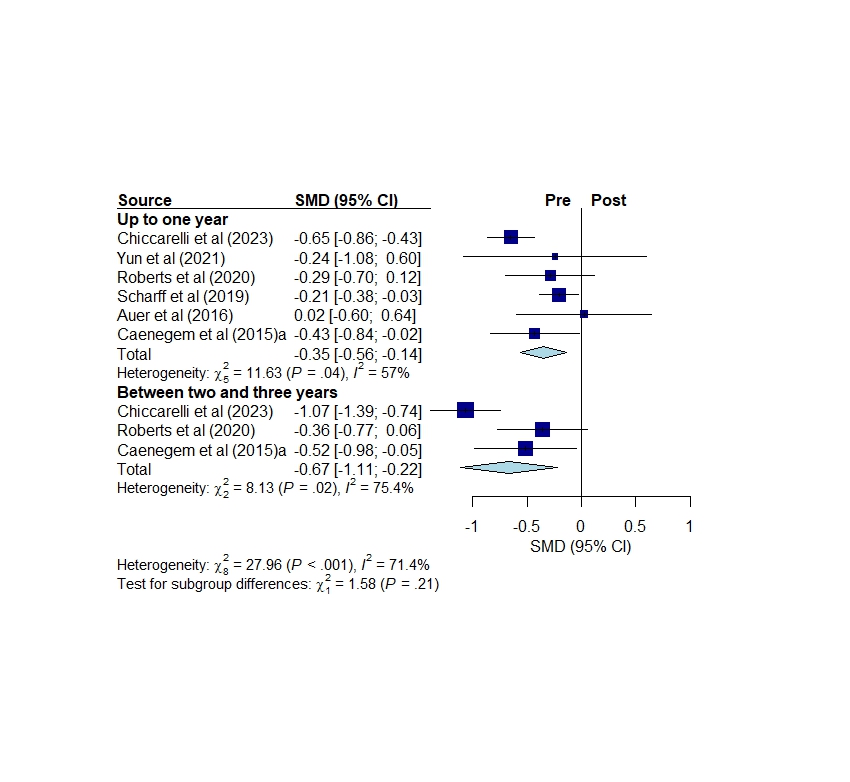

Figure S13. Forest plot: Upper-body strength pre vs. post hormone therapy in hormone-naïve transgender women.

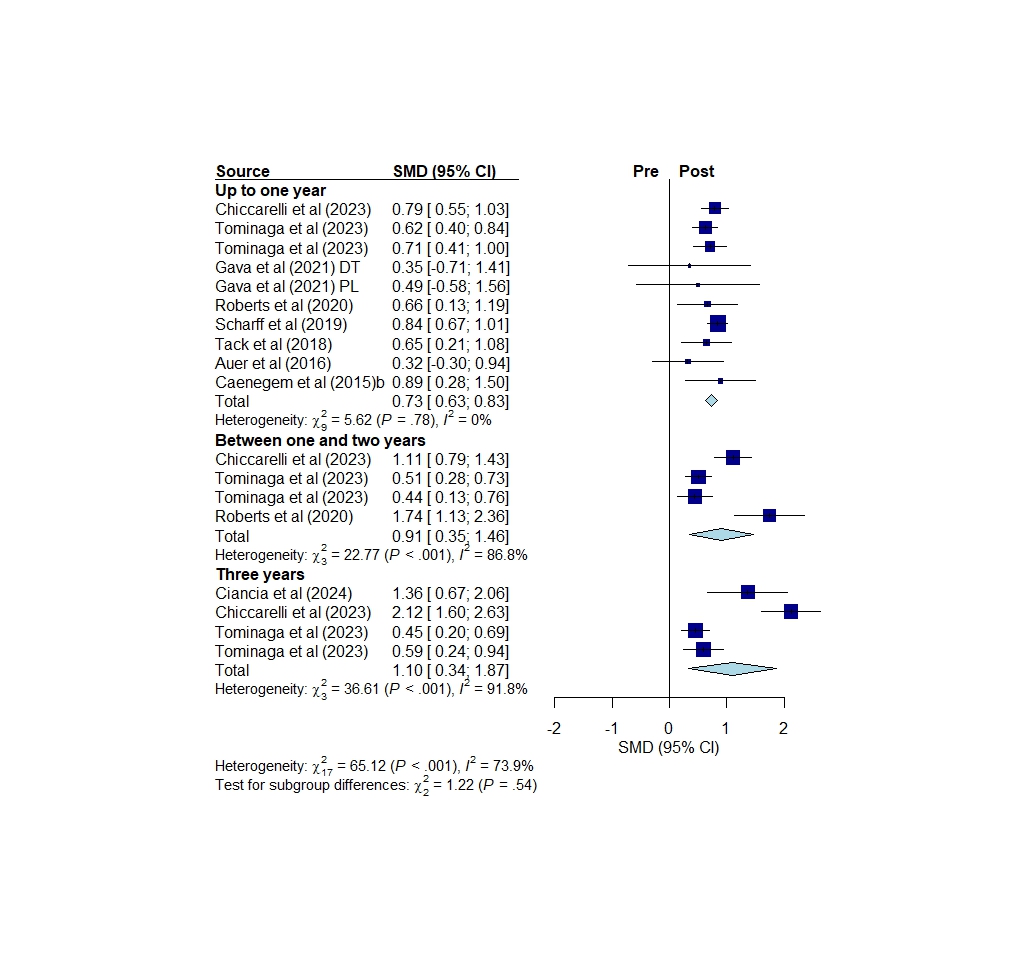

Figure S14. Forest plot: Upper-body strength pre vs. post hormone therapy in transgender men (all studies).

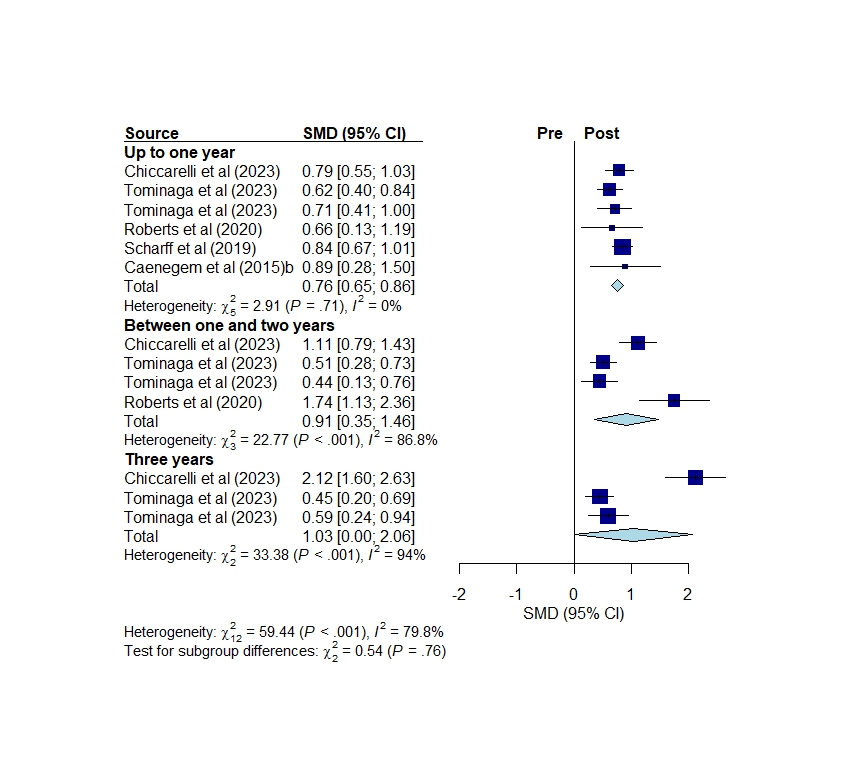

Figure S15. Forest plot: Upper-body strength pre vs. post hormone therapy in hormone-naïve transgender men.

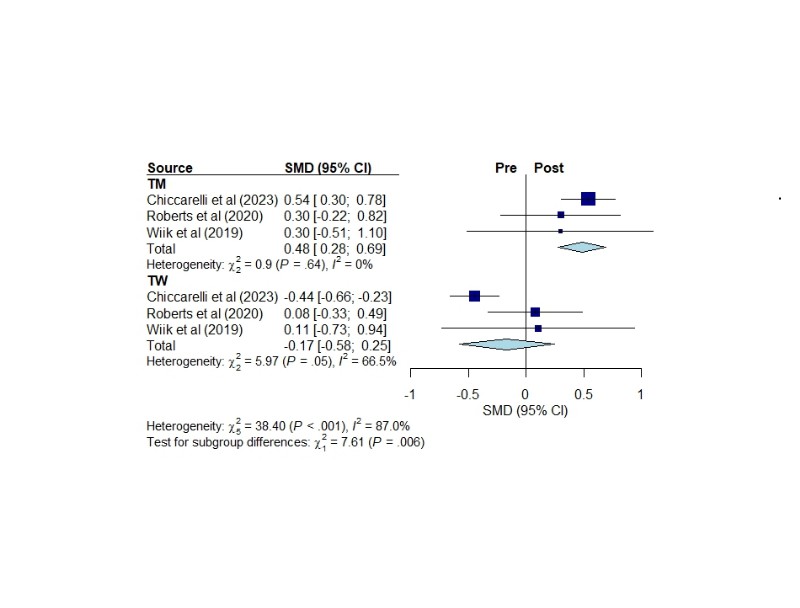

Figure S16. Forest plot: Lower-body strength pre vs. post hormone therapy in hormone-naïve transgender men (TM) and women (TW).

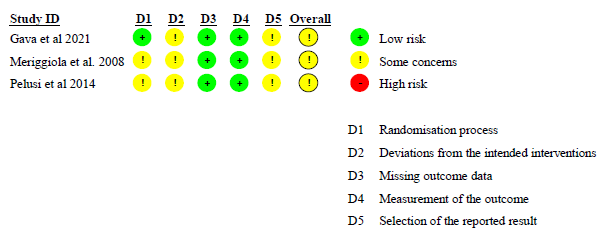

Figure S17. Quality assessment of randomized clinical trials using ROB2 (Risk Of Bias 2).
